# Leveraging haplotype information in heritability estimation and polygenic prediction

**DOI:** 10.1101/2024.04.30.24306654

**Authors:** Jonas Meisner, Michael Eriksen Benros, Simon Rasmussen

## Abstract

Polygenic prediction has yet to make a major clinical breakthrough in precision medicine and psychiatry, where the application of polygenic risk scores are expected to improve clinical decision-making. Most widely used approaches for estimating polygenic risk scores are based on summary statistics from external large-scale genome-wide association studies, which relies on assumptions of matching data distributions. This may hinder the impact of polygenic risk scores in modern diverse populations due to small differences in genetic architectures. Reference-free estimators of polygenic scores are instead based on genomic best linear unbiased predictions and models the population of interest directly. We introduce a framework, named `hapla`, with a novel algorithm for clustering haplotypes in phased genotype data to estimate heritability and perform reference-free polygenic prediction in complex traits. We utilize inferred haplotype clusters to compute accurate SNP heritability estimates and polygenic scores in a simulation study and the iPSYCH2012 case-cohort for depression disorders and schizophrenia. We demonstrate that our haplotype-based approach robustly outperforms standard genotype-based approaches, which can help pave the way for polygenic risk scores in the future of precision medicine and psychiatry. `hapla` is freely available at https://github.com/Rosemeis/hapla.

## Introduction

Polygenic prediction has become a major field of interest due to its potential impact and role in personalized medicine using estimated polygenic risk scores (PRSs) [51]. A PRS is a summarization of an individual’s genetic predisposition for a given disease or trait and is in its simplest form a weighted sum of risk alleles usually derived from large-scale genome-wide association studies (GWAS). However, most common diseases are complex traits and have thousands of risk-associated single-nucleotide polymorphisms (SNPs) with very small effect sizes, limiting the breakthroughs for the clinical utility of PRSs in personalized medicine. Early large-scale quantitative genetics research has additionally been biased towards individuals of European ancestry, further limiting the clinical utility of PRSs in modern diverse populations and the majority of the global population [17].

Polygenic prediction is also of great interest to precision psychiatry, where the complex etiologies of psychiatric disorders are yet to be untangled, and PRSs have the potential to aid in the clinical decision-making and individualized treatment for patients in the future [36, 16]. Major depressive disorder (MDD) is a disabling common illness and an increasing global health concern as it contributes substantially to the growing disease burden [24], whereas schizophrenia (SCZ) remains a substantial burden to society even with a low population prevalence of approximately 1% [8, 40]. Severe mental disorders therefore call for action to further efforts in understanding the genetic contributions in their etiologies. MDD and SCZ are both highly polygenic traits with 243 identified risk loci from a large-scale meta-analysis spanning multiple cohorts [3] for MDD, and 287 distinct loci for SCZ as well from a large-scale meta-analysis [52].

The two main approaches for performing polygenic prediction are denoted reference-based and reference-free, also referred to as summary statistics based and individual-level data based, respectively. Recent work shows that the combination of the two approaches can lead to increased predictive performance for complex traits [2]. The reference-based approach is based on summary statistics from an external large-scale GWAS, where the marginal regression coefficients of the SNPs are used to construct a PRS estimator [56, 10, 28]. Unfortunately, the reliance on an external large-scale GWAS has shown the now well-known portability problem in PRS estimators [31]. Issues arise due to assumptions of similarity in the generative phenotype process between the reference population and the target population including their linkage disequilibrium (LD) patterns leading to restrictions in the SNP set used for polygenic prediction [20, 9, 37, 17]. The genetic distance between the reference population and the target population has been shown to be directly correlated with the portability issues of a constructed PRS estimator [12]. Even for homogeneous ancestries, it has been shown that reference-based PRS estimators are still strongly influenced by the external GWAS, as PRSs can correlate with uncorrected population structure [48, 44, 46].

Reference-free polygenic prediction does not rely on pre-estimated summary statistics of an external GWAS but is instead based on the joint modeling of SNPs in the target population using a linear mixed model (LMM) [61], thus accounting for LD between SNPs [63]. LMMs are the state-of-the-art approach to account for population structure and cryptic relatedness in modern GWAS methods [18, 27]. The LMM frameworks estimate SNP heritability of a trait based on genome-based restricted maximum likelihood (GREML) given an estimated genome-wide relationship matrix (GRM) [62]. The inferred SNP heritability is implicitly used in the prediction of a polygenic score as well, also known as the best linear unbiased prediction (BLUP) [45]. However, recently it has been shown that naive BLUPs overfit the genetic effect by modeling non-genetic effects as well, and the study introduces a well-performing leave-one-out cross-validated BLUP (cvBLUP) estimator [33].

Haplotype-based methods have received greater interest in recent years due to their increased power in detecting fine-scale population structure [22, 34], local ancestry inference [30], heritability estimation and GWAS [5], and additionally, having increased accuracy in genomic prediction [54, 59] but they has yet to make an impact in precision medicine and psychiatry. These methods directly model LD patterns through haplotype information and should therefore be able to tag unobserved causal SNPs, which are poorly captured by an SNP array or imputed genotype data, due to the unobserved SNPs likely being in identity-by-state for long haplotypes being in identity-by-state [41]. Multiple approaches including local ancestry tracts inferred from haplotypes have also been developed to estimate SNP heritability [65] and construct reference-based PRS estimators [57].

In this work, we introduce a novel haplotype-based framework, `hapla`, which infers haplotype clusters in windows along the genome in a fast and parallelized approach. We utilize haplotype cluster assignments to construct a GRM, estimate heritability, and perform reference-free polygenic prediction in complex traits using cvBLUPs outperforming standard genotype-based approaches. We show and evaluate its performance in a simulation study and hereafter showcase its capabilities in heritability estimation and polygenic prediction of depression disorders and schizophrenia in the large Danish Integrative Psychiatric Research Consortium (iPSYCH) 2012 case-cohort [39]. The gain from leveraging haplotype information efficiently has the potential to advance precision medicine and psychiatry in modern diverse populations.

## Methods

### hapla framework

We here describe our `hapla` framework. We denote **X** ∈{0, 1} ^2*N×M*^ as the haplotype matrix of a chromosome for a given dataset of phased genotypes, consisting of 2*N* haplotypes and *M* diallelic SNPs. The haplotype matrix is assumed to be ordered such that two adjacent rows correspond to the maternal and paternal haplotypes of a single individual resulting in a total of *N* individuals. We split the chromosome into *W* genomic windows of a fixed size that can be overlapping or non-overlapping. For the sake of simple mathematical notation, we define windows to be non-overlapping and to have a fixed length of *B* SNPs such that **X** = [**X**^(1)^, …, **X**^(*W*)^] with **X**^(*w*)^ ∈{0, 1} ^2*N×B*^ for *w* = 1, …, *W*. The objective is to cluster haplotypes independently in each genomic window into a set of *K* haplotype clusters, where the number of defined clusters can vary between the different windows to account for non-uniform genetic diversity across the chromosome. Hereafter, we use the haplotype cluster assignments to estimate heritability and perform reference-free polygenic prediction.

#### Clustering algorithm

The haplotype clustering algorithm is applied to each window along a given chromosome independently. We have adopted a nonparametric clustering algorithm based on DP-Means [21] and its recently proposed extension with a delayed cluster creation process, PDC-DP-Means [11]. The algorithms can be seen as generalizations of the *K*-Means algorithm, where *K*, the number of clusters, is unknown and inferred from the data. However, instead of minimizing the Euclidean distance between haplotypes and cluster centroids, we propose a delayed cluster creation algorithm based on minimizing the Manhattan distance between haplotypes and cluster medians [6]. We denote the algorithm as PDC-DP-Medians using similar terminology. The Manhattan distance is an appealing choice for binary genetic data (Hamming distance), as it can be viewed as the number of mutations between a haplotype and a cluster median, thus providing a natural evolutionary perspective to the constructed cluster medians. The cluster medians will here act as haplotype representatives of the inferred haplotype clusters. Our haplotype clustering approach (PDC-DP-Medians) is further detailed in Algorithm 1.

##### Algorithm 1

PDC-DP-Medians

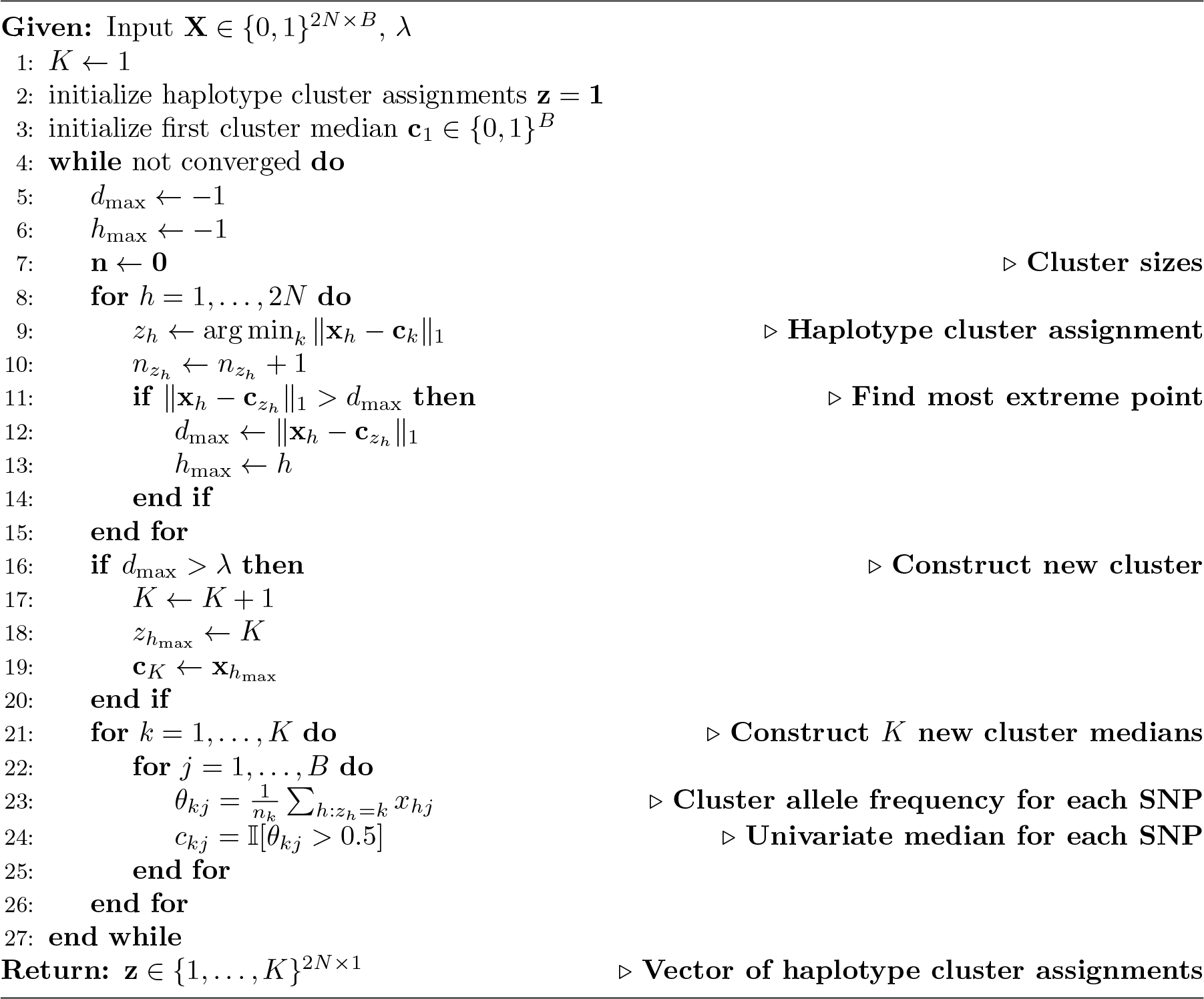

The following definitions will be for a single genomic window. The cost function of PDC-DP-Medians for *K* haplotype clusters is defined as:

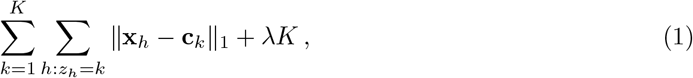

where **x**_*h*_ ∈ {0, 1}^*B*^ is the *h*-th haplotype, **c**_*k*_ ∈ {0, 1}^*B*^ is the *k*-th cluster median, *z*_*h*_ ∈ {1, …, *K*} is the haplotype cluster assignment of the *h*-th haplotype for *h* = 1, …, 2*N* and ∥ · ∥_1_ being the *L*_1_-norm. The added penalty term penalizes the creation of clusters, and a new cluster will be generated if the Manhattan distance exceeds the specified *λ* parameter. We utilize the delayed cluster creation process introduced in PDC-DP-Means, where only one new cluster can be created per epoch, which allows for a fast parallelized implementation of the algorithm.

For extending the median to multiple dimensions, we use the marginal median to define cluster medians, where each component in the multivariate median is defined as a simple univariate median, and it is thus fast to compute for binary genetic data. An example for the *j*-th SNP in **c**_*k*_:

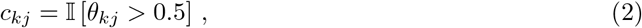

with 𝕀 [·] being the indicator function, 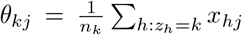 being the mean of the haplotypes assigned to the *k*-th cluster for SNP *j* and *n*_*k*_ the number of haplotypes assigned to cluster *k*. We initialize the algorithm with *K* = 1 and the first cluster median **c**_1_ being the marginal median considering all haplotypes such that *z*_*h*_ = 1 for *h* = 1, …, 2*N*. The next cluster creation step after initialization is then defined as finding the haplotype with the maximum distance to its assigned cluster, and if the distance exceeds *λ*, the corresponding haplotype is set as a new cluster. The process is iterated until the haplotype cluster assignment step converges, and we have set *λ* = 0.1 for all analyses in our study. The computational complexity for performing haplotype clustering in a single window will be 𝒪 (*NBK*). A simple visual depiction of our clustering algorithm is displayed in Figure 1.

**Figure 1:**
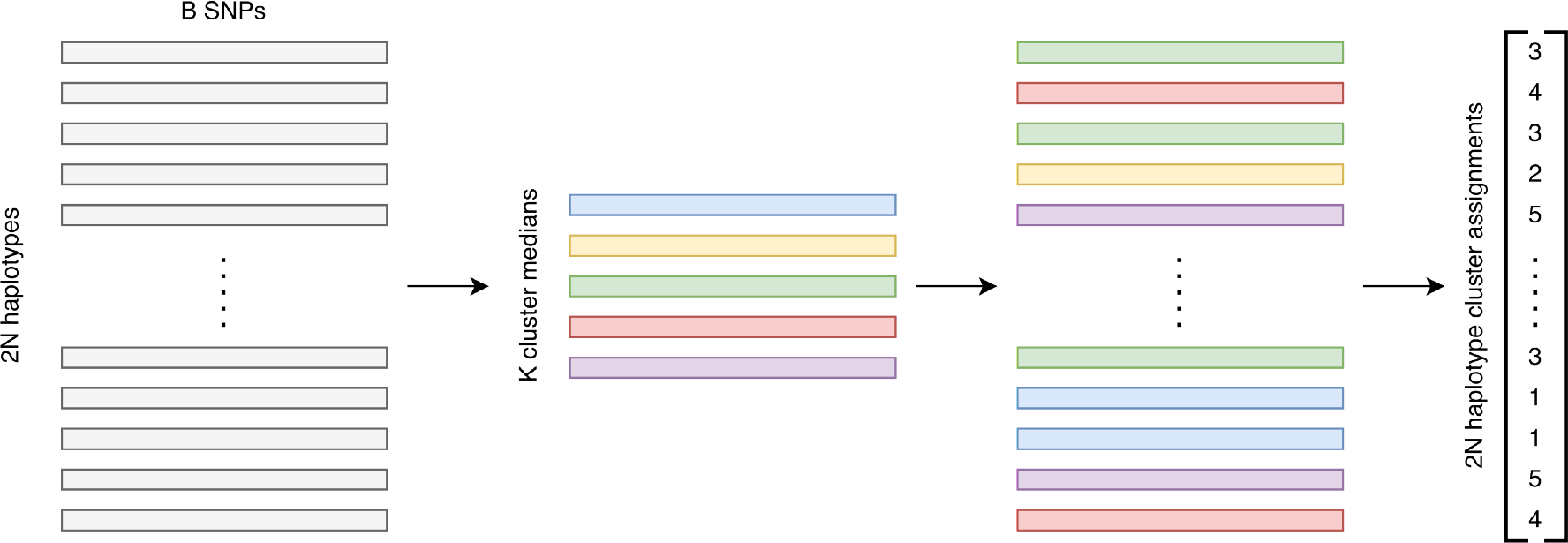
PDC-DP-Medians in the *w*-th genomic window of 2*N* haplotypes and *B* SNPs, **X**^(*w*)^. The haplotypes are assigned to the closest of the *K* inferred cluster medians, **c**^(*w*)^, by minimizing the Manhattan distance. The haplotype cluster assignment vector, **z**^(*w*)^, on the right side is the simplified output of the clustering algorithm.

When a window size of *B* = 1 is chosen, our algorithm naturally collapses to single SNP clusters representing the two alleles. We exclude rare haplotype clusters below a frequency threshold of *δ* = 0.01 in an iterative post-hoc re-clustering procedure, where haplotypes belonging to an excluded cluster will be reassigned to the next best cluster. We describe the procedure in Algorithm S1 in the supplementary material. This step increases the computational runtime of our algorithm but ensures that all haplotypes are represented by and assigned to valid clusters, thus retaining information. *λ* and *δ* can be seen as smoothing hyperparameters that affect granularity, as they govern how many haplotype clusters will be generated and retained, respectively.

#### Genome-wide relationship matrix

The haplotype cluster assignment of a single haplotype *h* in window *w* is one-hot encoded such that 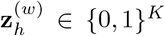, with the *k*-th entry as 1 and 0 otherwise for 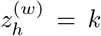. We define a new matrix **A**^(*w*)^ ∈ {0, 1, 2}^*N×K*^, which represents the aggregated haplotype cluster assignments for each individual, where 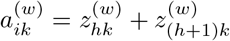 for the *i*-th individual with its maternal and paternal haplotypes (*h* and *h*+1) in the *w*-th window, for *k* = 1, …, *K*. We will refer to **A** = [**A**^(1)^, …, **A**^(*W*)^] as the full haplotype cluster allele matrix, which can span multiple chromosomes. Thereby in comparison to standard genetic data, we have changed the input unit from minor allele counts of SNPs (genotypes) to haplotype cluster allele counts. Note that each genomic window *w* has had an assumed fixed number of *K* haplotype clusters for simple notation, however in practice, *K* will vary in each window.

We define a general approach for estimating the genome-wide relationship matrix (GRM), **R** ∈ℝ^*N×N*^, with our haplotype cluster allele matrix. The pairwise entry of individual *i* and *j* is defined as follows:

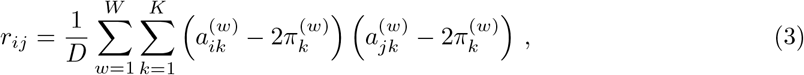

where 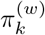 represents the haplotype cluster allele frequency of the *k*-th haplotype cluster in window *w* and 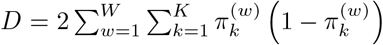 scales **R** to be analogous to the numerator relationship matrix [55]. We further perform data and Gower centering on **R** to obtain accurate heritability estimates [66] in the following approach.

Reference-free polygenic prediction is based on the best linear unbiased prediction (BLUP) from a linear mixed model (LMM), using the GRM estimated above, such that a phenotype **y** ∈ ℝ^*N*^ is modeled as a mixture of fixed and random effects using all SNPs, or in our case haplotype cluster alleles, jointly:

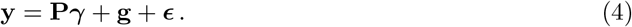

Here **P** ∈ ℝ^*N×Q*^ is a matrix of *Q* covariates, including inferred eigenvectors, with ***γ*** as the corresponding fixed effects, **g** ∈ ℝ^*N*^ is a vector of the genetic effects, or polygenic scores, of the individuals with 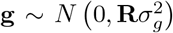 and ***ϵ*** ∈ ℝ^*N*^ is a vector of residual effects with 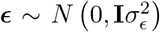. The linear mixed model can be extended to include genetic effects from multiple GRMs [64, 49] that each models different partitions of the data. The phenotype will therefore be modeled as

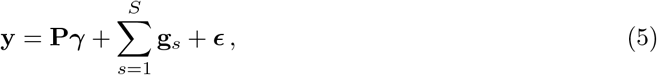

using *S* different GRMs with 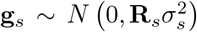. For our haplotype-cluster approach, it will be GRMs estimated from haplotype clusters inferred from different window sizes. The variance components are estimated using genome-based restricted maximum likelihood (GREML), and we define 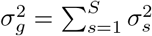.

#### Polygenic prediction

Reference-free polygenic prediction is implicitly performed using the GREML approach above to provide polygenic predictions in the target population with 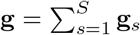, where **g**_*s*_ represents the predictions from the *s*-th partition. Importantly, we compute leave-one-out cross validated genetic predictions (cvBLUP) to avoid overfitting and biased estimates of the polygenic scores [33], which allows for direct and useful predictions in the target population. We use `GCTA` (v1.94.1) to estimate the variance components for SNP heritability and cvBLUPs using flags `--reml`, `--reml-no-constrain`, `--cvblup` and `--mgrm` to include multiple GRMs.

Reference-based polygenic prediction is estimated using external genome-wide association study (GWAS) summary statistics. We use `PRSice` (v2.3.5) [10] to estimate polygenic risk scores based on a clumping and thresholding (C+T) approach using default parameters that optimizes *R*^2^.

#### Implementation details

We have implemented our method in a fast and multithreaded command-line software, `hapla`, which is written in Cython (v3.0) and Python (v3.11). The software takes phased genotype data as input in VCF/BCF format using the `cyvcf2` (v0.30) Python library [38], which is read into `NumPy` (v1.26) arrays [14] for fast array manipulation. The phased genotype data is read into 1-bit arrays such that the memory requirement is approximately *NM/*4 bytes, which makes the haplotype clustering in the `hapla` software scalable to modern biobank sample sizes.

The output files of our software are either in binary `NumPy` format, binary `PLINK` format or binary `GCTA` format for fast and easy integration into existing pipelines. The software is open source and freely available at https://github.com/Rosemeis/hapla. All results generated in this study is based on `hapla` v0.6.

### Simulated data

We simulate human genetic data based on a two populations out-of-Africa demographic model (‘OutOfAfrica 2T12’) [50, 13] in the `stdpopsim` (v0.2) library [1] using the `msprime` (v1.2.0) simulation engine [19]. The simulated data is used to evaluate `hapla` in estimating heritability and performing polygenic prediction in a realistic scenario including populations of different genetic architectures. The demographic model is visualized in Figure S1, and we simulate a total of 20,000 individuals with 10,000 from each of the two populations of different ancestry, African (AFR) and European (EUR). A chromosomal segment of 100Mb is simulated using a constant recombination rate of 1.28 *×* 10^−8^ and mutation rate of 2.36 *×*10^−8^. We split the data into three subsets, AFR (*N* = 10,000), EUR (*N* = 10,000) and ALL (*N* = 20,000), where ALL represents a structured population containing all individuals. After minor allele frequency (MAF) filtering at a threshold of 0.01, we have 425,903 SNPs, 223,407 SNPs and 388,746 SNPs, respectively, in the three subsets emulating whole-genome sequencing data with full information.

We create downsampled datasets of the three subsets, where we downsample the SNP set by a factor of 10 to remove information that would potentially not be available in real datasets due to imperfect imputation or relying on SNP array data. We create 10 equally sized MAF bins from 0.01 to 0.5 and sample uniformly from each, which emulates a scenario where we have proportionally more frequent SNPs similar to SNP array data or imputed data in comparison to sequencing data. The downsampled datasets therefore only include 42,600, 22,350 and 38,880 SNPs for AFR, EUR and ALL, respectively.

Additionally, we create linkage disequilibrium (LD) pruned subsets of all mentioned datasets to evaluate a standard genotype-based method in which LD pruning has been performed. LD pruning is a common procedure to reduce the computational burden in LMMs and principal component analysis (PCA) [26]. It removes correlated SNPs in close proximity [32], and we perform standard LD pruning using `PLINK` (v2.00a) [7] with flag ‘`--indep-pairwise 50 10 0.5`’.

A complete overview of the generated simulated datasets is listed in Table S1.

#### Phenotype simulations

We simulate phenotypes of complex traits in the simulated data for three different scenarios. We follow the simple approach in [31], where we randomly sample *M*_*c*_ = 1,000 causal variants. Causal effects are assumed to be independent and sampled from a Gaussian distribution, 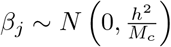, for *j* = 1, …, *M*_*c*_, where *h*^2^ is the narrow-sense heritability. The genetic liability of the trait for individual *i* is defined as an additive model of the causal variants such that

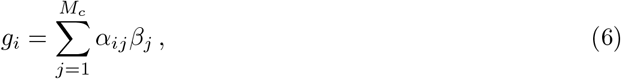

with *α*_*ij*_ being the genotype at the *j*-th causal variant. Environmental noise is sampled from a Gaussian distribution, *ϵ*_*i*_ ∼ *N* (0, 1 − *h*^2^), and the phenotype of an individual *i* is then generated by 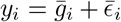. Here 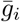 and 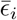 represent scaled versions of *g*_*i*_ and *ϵ*_*i*_, which ensure that their variances are *h*^2^ and 1 − *h*^2^, respectively. We simulate SNP-based phenotypes using two different *h*^2^, which are 0.8 (Scenario 1) and 0.2 (Scenario 2). We also simulate phenotypes of quantitative traits using haplotype cluster alleles for *h*^2^ = 0.8 to demonstrate that our approach using haplotype clusters is unbiased (Scenario 3). We use haplotype clusters inferred from *B* = 8.

Each simulation scenario is replicated 10 times to account for sampling bias, and the results are reported with mean and standard deviation across the 10 replications.

### iPSYCH2012 case-cohort

The Integrative Psychiatric Research Consortium (iPSYCH) 2012 is a Danish population-based case-cohort of individuals with mental disorders and population controls [39]. The iPSYCH2012 case-cohort consists of selected Danish individuals born between 1981 and 2005, where dry blood spots have been taken at birth and have been genotyped on a Infininium PsychChip v1.0 array containing a large proportion of SNPs associated with psychiatric disorders. The population controls are part of a large random sample of individuals from the Danish population and can therefore have any of the disorders by chance. Following the extensive sample and SNP quality control including haplotype estimation performed in the iPSYCH2012 case-cohort [4], we utilize the generated phased imputed genotype data to estimate heritability and perform polygenic prediction for major depressive disorder (MDD), recurrent depressive disorder (REC) and schizophrenia (SCZ) using our inferred haplotype clusters. MDD is here defined by the 10th revision of International Classification of Disease (ICD-10) [58] codes F32 and F33, whereas REC is a subtype of MDD (F33 only) and SCZ is defined by the ICD-10 code F20. All three disorders are analyzed independently, and population controls with a diagnosis of the given disorder in the Danish Psychiatric Central Research Register [35] are then classified as cases in the respective analyses, and the population controls will therefore differ between the disorders. We only consider individuals that are active, alive and residents in Denmark at the end of 2016 from where we also calculate their respective age.

We filter out SNPs deviating from Hardy-Weinberg equilibrium (HWE) with *p <* 1.0 *×* 10^−6^ as well as using an imputation quality filter of DR2 ≤ 0.95 to estimate kinship coefficients using KING-robust [29], implemented in `PLINK`, while pooling population controls and cases from all three disorders. We remove related individuals up to second-degree (0.0884), while keeping individuals in the related pairs that maximize the overall sample size. Using these definitions, we have 20,620 cases with a MDD diagnosis, 7,128 cases with a REC diagnosis and 3,211 cases with a SCZ diagnosis, and 25,228, 25,840 and 25,994 population controls in the three datasets, respectively. Hereafter we filter out SNPs with MAF *<* 0.01 and we provide a full overview of the number of SNPs in the three datasets in Table S3. We use a population life-time disease prevalence of 0.12285 for MDD, 0.05700 for REC and 0.01745 for SCZ [40, 47] to convert SNP heritability estimates on the observed scale to the liability scale [23]. We infer the top 20 principal components using the `PCAone` (v0.4.2) software [25], which is used to correct for population structure in downstream analyses.

## Results

### Heritability estimation and polygenic prediction in simulation study

We had generated genetic data of a 100Mb chromosomal segment for a total of 20,000 individuals from two different populations in an out-of-Africa demographic model (AFR and EUR) to evaluate the capabilities of `hapla` in heritability estimation and polygenic prediction. We also evaluated the combined data, denoted ALL, to investigate the effect of modelling a structured population. We simulated three different phenotype scenarios sampling 1000 causal loci, each replicated 10 times, where Scenario 1 and 2 were SNP-based assuming *h*^2^ = 0.8 and *h*^2^ = 0.2, respectively, and Scenario 3 was haplotype cluster-based assuming *h*^2^ = 0.8, in each of the three datasets. Additionally, we generated datasets with downsampled SNP sets to emulate loss of information due to for example imperfect imputation or using SNP arrays, which we evaluated as well for all scenarios. We reported the SNP heritability estimates and the predictive performance of the estimated polygenic scores (cvBLUPs), which were evaluated using a standard *R*^2^ metric with the phenotypes.

#### Window sizes in hapla and multiple genetic effects

The window size used in `hapla` might affect the haplotype information extracted, and we therefore investigated its performance using different window sizes, as well as a joint approach with multiple genetic effects, to find an optimal modelling procedure. We inferred haplotype clusters in overlapping windows across the simulated chromosomal segment of different SNP lengths, *B* = {1, 8, 16, 32} (Table S2), where *B* = 1 represented standard SNP information within our framework. We clustered the haplotypes in under 25 minutes (using 8 threads) for the different window sizes in all datasets showcasing the speed of our clustering algorithm (Figure S2).

We therefore evaluated the window sizes independently, but also jointly, where we estimated multiple genetic effects by including the computed GRMs for each of the window sizes, which we referred to as “Multi” in the following results. The results of the different window sizes were shown in Figure S3 and Table S5 for Scenario 1. When evaluating the window sizes independently using the full datasets, the heritability estimates were very consistent around 0.8 across the different subsets, however, the predictive performance decreased for larger window sizes in comparison to *B* = 1, of which the phenotypes were simulated from. A similar pattern of consistent heritability estimates was observed for the downsampled datasets except for *B* = 1, which heavily underestimated the heritability in AFR with 0.6702, however, we saw again a general decrease in the predictive performance for increasing window sizes. In Scenario 2 (Figure S4 and Table S6), the phenotypes were simulated using *h*^2^ = 0.2. We observed the same overall consistent heritability estimates across the different window sizes using both full and downsampled datasets, except for *B* = 1 using the downsampled datasets, which again underestimated the heritability in all three subsets with 0.1494, 0.1781 and 0.1700 for AFR, EUR and ALL, respectively. The genetic predictions followed the same trend as in Scenario 1, where the predictive performance decreased for larger window sizes. Lastly, Scenario 3 produced almost identical results to Scenario 1, though with an overall reduced performance across all combinations for both the full and downsampled datasets (Figure S5 and Table S7). Here *B* = 8, of which the phenotypes were simulated from, had the overall best performance using the full datasets as expected.

Lastly, we modeled the genetic effects from each GRM jointly, and we observed that it performed almost identically to *B* = 1 in Scenario 1 and 2 and *B* = 8 in Scenario 3 using the full datasets, which were also separate partitions in the Multi approach. It outperformed every other configuration in all three scenarios using the downsampled datasets in terms of predictive performance of polygenic scores, and it were almost matching the best performing configuration in heritability estimations, which was most often itself. This indicated that the different window sizes captured different haplotype information, such that the Multi approach could take advantage of all partitions and retained overall high accuracy in heritability estimates and predictive performance of polygenic scores, and thus, we would only consider the Multi approach from this point on.

#### Comparison of hapla to genotype-based approaches

We compared the results of the `hapla` approach using multiple genetic effects from different window sizes to two genotype-based GRMs computed in `GCTA` to demonstrate the gain of using haplotype information in heritability estimation and polygenic prediction. Here one was estimated in a standard procedure, referred to as “`GCTA`”, and the other was based on LD pruned genotype datasets, referred to as “Pruned”, which was a common procedure to reduce computational burdens.

In Scenario 1 (*h*^2^ = 0.8), we immediately observed that `hapla` was outperforming the other two approaches in all of the three subsets for both the full and the downsampled datasets based on predictive performance of its polygenic scores (Figure 2 and Table S8). The predictive performance of the standard GRM in `GCTA` was relatively close to `hapla` in the all three subsets, even though the heritability estimates were much less accurate. Our haplotype-cluster approach had relative increases in *R*^2^ of 6.2%, 3.8% and 5.4% in the full data for AFR, EUR, and ALL, respectively, and 4.6%, 2.7% and 3.3% in the downsampled data in comparison to `GCTA`. The heritability estimates of `hapla` were consistently always more accurate, except for ALL using the full dataset, where it slightly overestimated the heritability with 0.8207 in comparison to 0.8117 and 0.8043 for `GCTA` and pruned, respectively. Meanwhile in the downsampled datasets, `hapla` were almost on par with the heritability estimates from the full data with 0.7811, 0.7722 and 0.7986 for AFR, EUR and all, respectively, whereas the genotype-based approaches in general severely underestimated the SNP heritability.

**Figure 2:**
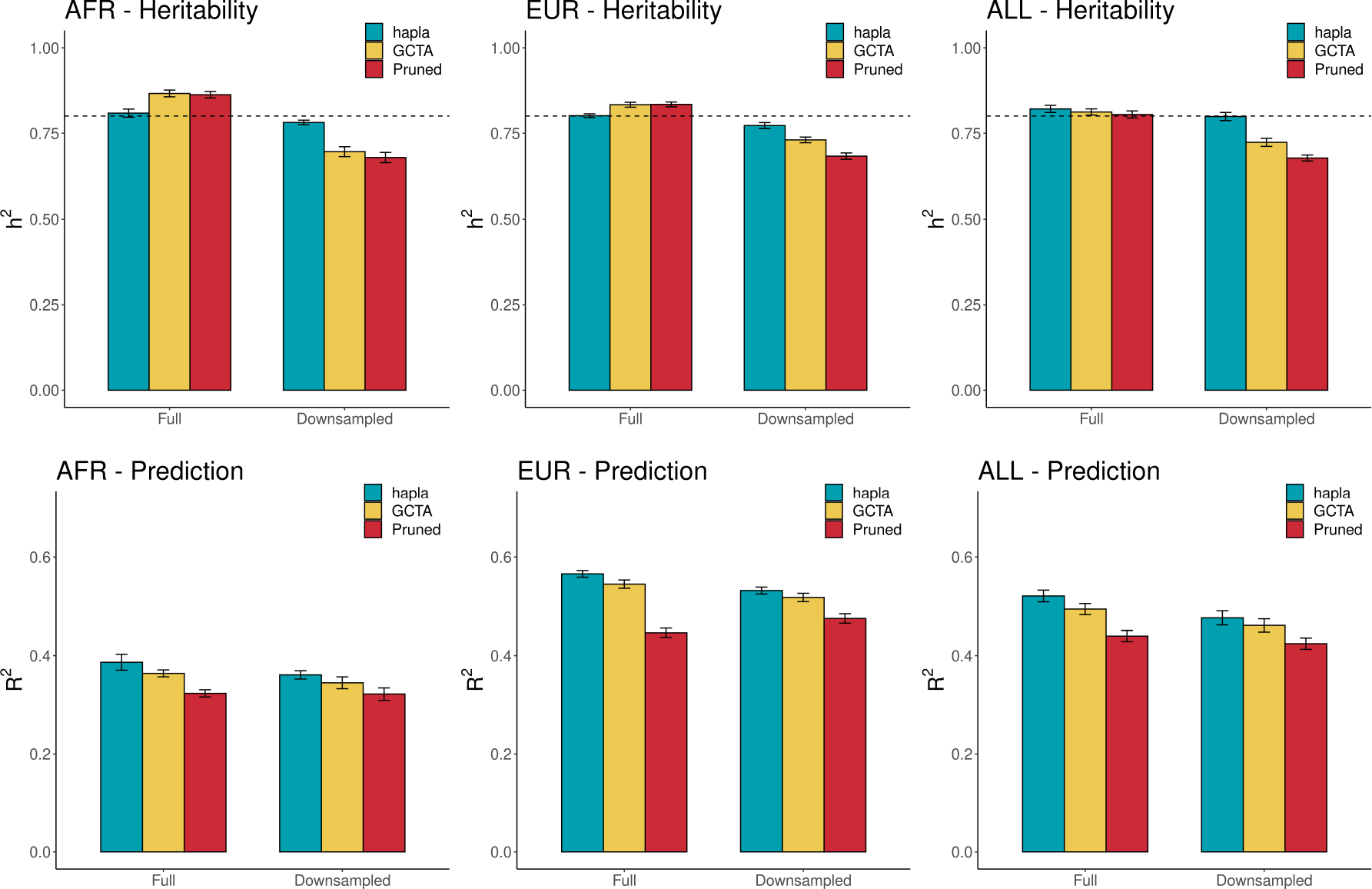
Heritability estimation and polygenic prediction in Scenario 1 across 10 phenotype simulations. Top row shows the SNP heritability estimates using the different GRMs across three subsets using either full or downsampled data, while the bottom row are the squared correlations between the cvBLUPs and the simulated phenotypes. The true simulated *h*^2^ = 0.8 is displayed with a dashed line and the error bars represent the standard deviation.

Next, we performed the same analyses in Scenario 2 with *h*^2^ = 0.2, and we similarly saw that `hapla` outperformed `GCTA` and Pruned in all scenarios based on both heritability estimation and polygenic prediction, except for the heritability estimate in ALL using the full dataset with 0.2077, 0.2089 and 0.2027 for `hapla`, `GCTA` and pruned, respectively (Figure S6 and Table S9). Interestingly, we observed a large overestimation of the SNP heritability for both `GCTA` and Pruned in AFR and EUR using the full dataset. The relative increases in *R*^2^ for `hapla` in the full data were 11.6%, 7.2% and 8.4% for AFR, EUR and ALL, respectively, and 3.3%, 4.0% and 2.3% in the downsampled data, when again compared to `GCTA`, thus showcasing larger gains from using haplotype information in the full data for Scenario 2.

In the last scenario for haplotype cluster-based phenotypes with *h*^2^ = 0.8, we observed identical results compared to Scenario 1, though with overall decreased performance for all approaches (Figure S7 and Table S10). However, the gap in predictive performance between `hapla` or `GCTA` did slightly increase in favour of `hapla` on average. We saw relative increases of 4.7%, 5.0% and 4.7% for AFR, EUR, and ALL, respectively, in the full data and 9.4%, 4.5% and 5.9% in the downsampled data. Overall we observed that LD pruning only affected results negatively in polygenic prediction across all tested scenarios and almost only produced worse heritability estimates in comparison to the standard genotype-based approach. `hapla` therefore in general produced more accurate and unbiased heritability estimates and more predictive polygenic scores in the simulated datasets in comparison to genotype-based approaches across multiple phenotype scenarios and diverse data subsets.

### Predicting mental disorders in the iPSYCH2012 case-cohort

Following the results of the simulation study, we inferred haplotype clusters and computed GRMs in the phased imputed genotype datasets of the iPSYCH2012 case-cohort using the same window sizes and modeled multiple genetic effects (Table S2). We evaluated three mental disorder categories, major depressive disorder (MDD), recurrent depressive disorder (REC) and schizophrenia (SCZ), which each consisted of 20,620, 7,128 and 3,211 cases, respectively. We included age, gender and the top 20 principal components as fixed-effects covariates in GREML and cvBLUP estimations, where we reported the log-likelihood, heritability estimates on both the observed and the liability scale, and the polygenic risk scores were evaluated using Nagelkerke’s pseudo 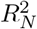 metric.

#### Heritability estimation and reference-free polygenic risk scores

Similarly to the simulation study, we compared `hapla` to standard genotype-based approaches in heritability estimation and predictive performance of reference-free polygenic risk scores. `hapla` had better predictive performance of 6.9%, 12.7% and 12.4% relative increases in 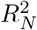 compared to the standard genotype-based approach in `GCTA` for MDD, REC and SCZ, respectively, and of 22.1%, 32.3% and 29.9% relative increases compared to the LD pruning approach (Figure 3 and Table 1). Similar to what we found in the simulation study, we expected heritability estimates of `hapla` to be more accurate and less biased, and we observed that `hapla` and `GCTA` produced relatively similar heritability estimates with 0.1448, 0.1113 and 01491 for MDD, REC and SCZ, respectively, using `hapla`, and 0.1442, 0.1070 and 0.1544 using the standard approach in `GCTA` on the liability scale. The heritability estimates of `hapla` were slightly larger for the depression disorders but lower for schizophrenia. As we observed in the simulation study, LD pruning only affected the performances negatively in polygenic prediction and overestimated heritability in comparison to the two other approaches. The log-likelihoods from GREML estimations using `hapla` were consistently the highest for all three mental disorders alongside predictive performance, thus further showcasing the gain from the inferred haplotype clusters.

**Table 1:**
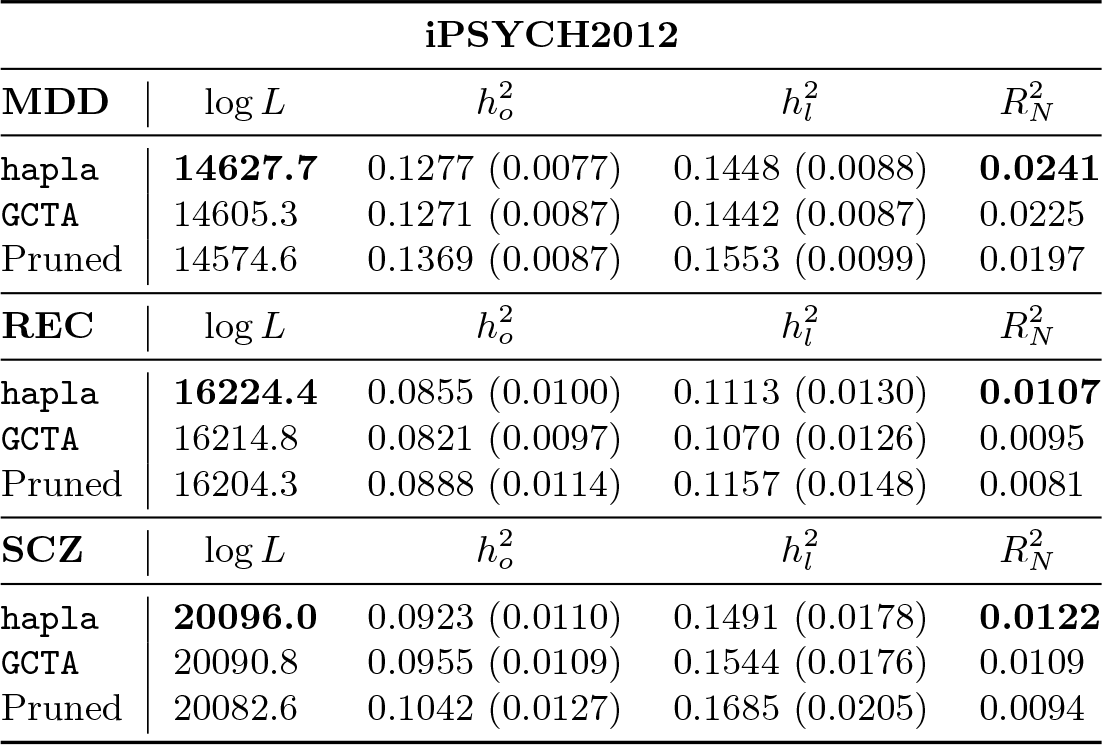
GREML-based log-likelihoods, estimates of SNP heritability and polygenic predictions of major depressive disorder (MDD), recurrent depressive disorder (REC) and schizophrenia (SCZ) in the imputed data of the iPSYCH2012 case-cohort. The SNP heritability estimates are converted to the liability scale assuming a population life-time prevalence of 0.12285, 0.05700 and 0.01745 for MDD, REC and SCZ, respectively. The standard errors of the heritability estimates are reported in parentheses. The polygenic risk scores from cvBLUP are evaluated based on Nagelkerke’s pseudo *R*^2^ measure with the highest values marked in bold as well as for the log-likelihood using GREML.

| iPSYCH2012 |  |  |  |  |
| --- | --- | --- | --- | --- |
| MDD | $\log L$ | $h_o^2$ | $h_l^2$ | $R_N^2$ |
| hapla | **14627.7** | 0.1277 (0.0077) | 0.1448 (0.0088) | **0.0241** |
| GCTA | 14605.3 | 0.1271 (0.0087) | 0.1442 (0.0087) | 0.0225 |
| Pruned | 14574.6 | 0.1369 (0.0087) | 0.1553 (0.0099) | 0.0197 |
| REC | $\log L$ | $h_o^2$ | $h_l^2$ | $R_N^2$ |
| hapla | **16224.4** | 0.0855 (0.0100) | 0.1113 (0.0130) | **0.0107** |
| GCTA | 16214.8 | 0.0821 (0.0097) | 0.1070 (0.0126) | 0.0095 |
| Pruned | 16204.3 | 0.0888 (0.0114) | 0.1157 (0.0148) | 0.0081 |
| SCZ | $\log L$ | $h_o^2$ | $h_l^2$ | $R_N^2$ |
| hapla | **20096.0** | 0.0923 (0.0110) | 0.1491 (0.0178) | **0.0122** |
| GCTA | 20090.8 | 0.0955 (0.0109) | 0.1544 (0.0176) | 0.0109 |
| Pruned | 20082.6 | 0.1042 (0.0127) | 0.1685 (0.0205) | 0.0094 |

**Figure 3:**
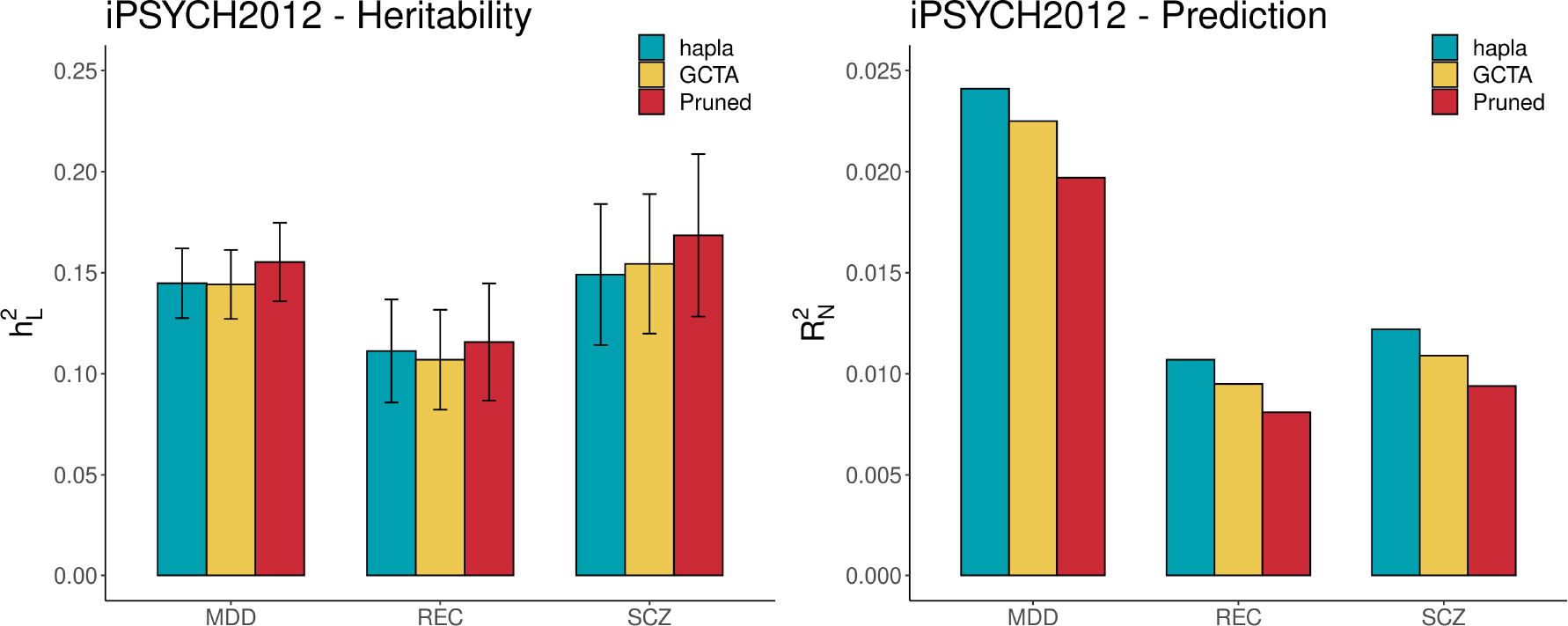
Heritability estimation and polygenic prediction in the iPSYCH2012 case-cohort. Left plot shows the SNP heritability estimates on the liability scale using the different GRMs across the three disorders, while the right plot shows the Nagelkerke’s pseudo *R*^2^ between the cvBLUPs and the binary outcomes. The error bars represent the 95% confidence interval for the heritability estimates.

#### Reference-based polygenic risk scores from summary statistics

We computed reference-based polygenic risk scores using a standard clumping and thresholding approach based on GWAS summary statistics from individuals of European ancestry in the Psychiatric GWAS Consortium (PGC) [15, 53] excluding iPSYCH case-cohorts for MDD and SCZ to evaluate against the reference-free approaches. The reference-based polygenic risk scores were based on ∼2.5x and ∼20x more cases than were available in the iPSYCH2012 case-cohort for MDD and SCZ, respectively (Table S11). However, their predictive performance was 0.0047 and 0.0121 for the disorders, and they did therefore not outperform `hapla` (0.0241 and 0.0122) or even the other genotype-based reference-free approaches for MDD.

We further computed correlations between the reference-based polygenic risk scores and the estimated principal components, which were used as fixed-effects covariates in all the reference-free estimations, for indications of uncorrected population structure in the GWASs. This clearly demonstrated that the reference-based polygenic risk scores correlated with structure in the iPSYCH2012 case-cohort captured by the principal components in comparison to `hapla` (Figure S8). The reference-based polygenic risk scores were highly correlated with PC1 (*r* = 0.2664) and PC6 (*r* = − 0.2024) for MDD, as well as for SCZ (*r* = 0.3513 and *r* = 0.4776, respectively), which could inflate their predictive performance due to modelling non-genetic effects through uncorrected population structure but still did not outperform `hapla`.

#### Predictive performance in individuals with at least one non-Danish born parent

Reference-based polygenic risk scores suffer from portability problems, and we investigated the predictive performances of all approaches in subsets of individuals in the iPSYCH2012 case-cohort, based on the country of birth of their parents, as we did not remove individuals due to ancestry or ethnicity in the reference-free approaches. We observed that all approaches had increased performances when focusing on individuals with both parents born in Denmark in comparison to evaluations on the full sample (Figure 4 and Table S12). The portability problem for reference-based polygenic scores were clearly shown for individuals with at least one parent not born in Denmark, where the predictive performance was severely impacted for MDD and SCZ. The reference-free polygenic risk scores were in general more robust, as there had also been no filtering based on ancestry or ethnicity in the GREML and cvBLUP estimations. `hapla` experienced the smallest change in predictive performance between the subsets of individuals with relative decreases in 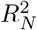 of 8.1%, 4.6% and 1.6% for MDD, REC and SCZ, respectively, compared to its best values, while outperforming all other approaches. This was in stark contrast to the reference-based polygenic risk scores that collapsed with relative decreases of 97.9% and 31.3% for MDD and SCZ, respectively. We therefore generated more predictive and robust polygenic risk scores for individuals in the iPSYCH2012 case-cohort using our haplotype cluster approach against both standard reference-free and reference-based approaches, and even when comparing to reference-based polygenic risk scores from very large sample sizes in individuals with both parents born in Denmark.

**Figure 4:**
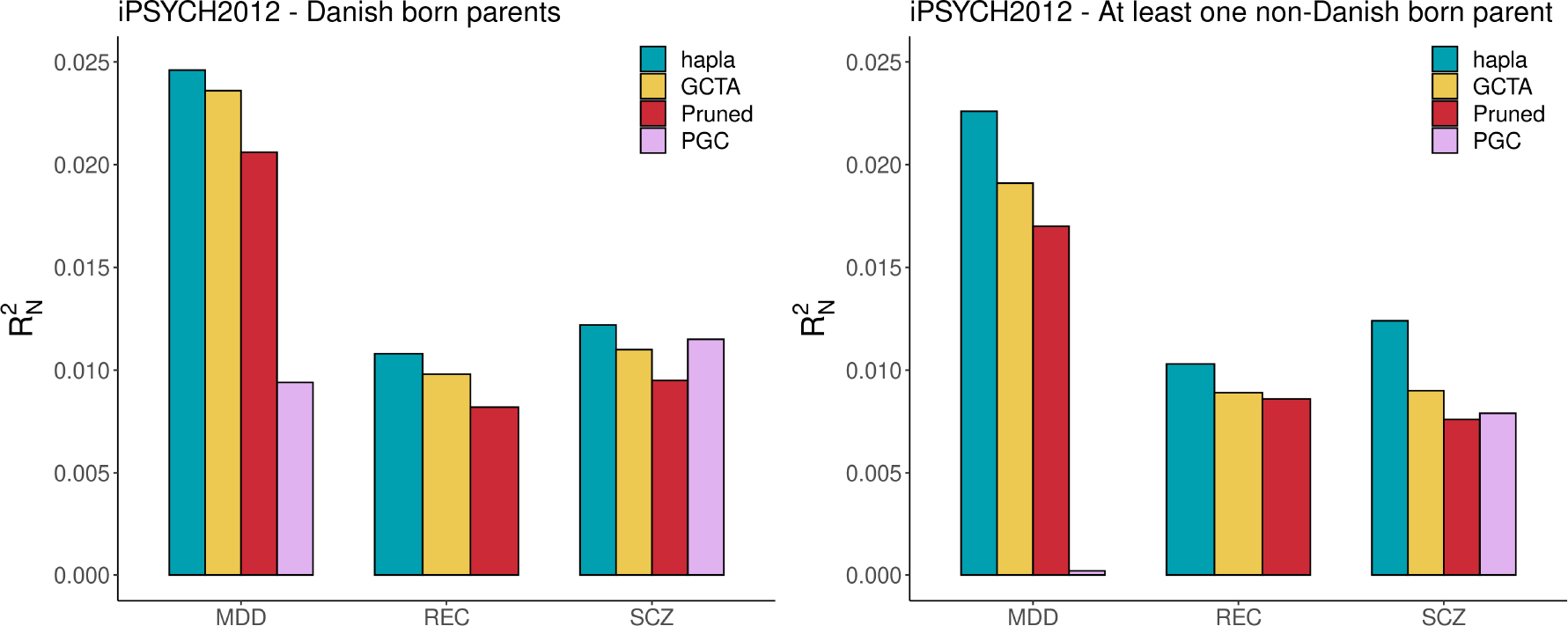
Predictive performance of polygenic risk scores in subsets of the iPSYCH2012 case-cohort. The left plot shows the performance on the subset of individuals with both parents being born in Denmark, while the right plot shows the performance on the subset of individuals with at least one parent born outside Denmark. “PGC” represents the reference-based polygenic risk scores from PGC GWAS summary statistics using `PRSice`.

## Discussion

We presented our framework, `hapla`, which performed window-based haplotype clustering in phased genotype data using our proposed PDC-DP-Medians algorithm. We explored the benefits of leveraging haplotype information in heritability estimation and reference-free polygenic prediction through the inferred haplotype clusters. In an extensive simulation study, we showcased the performance gain from our inferred haplotype clusters in multiple scenarios of diverse data subsets. `hapla` outperformed the widely used genotype-based GREML approach in `GCTA`, when evaluating the accuracy of SNP heritability estimates and the predictive performance of estimated polygenic scores in the simulated data. We hypothesize that this is due to an increased tagging ability of the inferred haplotype clusters. Another innovation in `hapla` is that we combine GRMs computed from haplotype clusters of different windows sizes and through this retain more haplotype information, leading to an increase in accuracy of both heritability estimates and polygenic scores. Strikingly, the gain from utilizing haplotype clusters was even more pronounced in the downsampled datasets that resemble common cases of imperfect imputation or SNP arrays. Here `hapla` showcased superior performance based on both heritability estimates and polygenic scores, which almost matched the genotype-based approach having full information. This is highly promising for genetic studies where sequencing data or accurate imputation are not available. Additionally, we show that even though LD pruning has become a common procedure in large-scale genetic studies in order to lower computational costs, we only observed that it was detrimental to performance and should therefore be avoided.

Due to the current reliance on external summary statistics for polygenic risk score estimation, it has become common practice to remove admixed and individuals of different ancestries in genetic studies that potentially affects relative large proportions of modern populations. When applying `hapla` to the iPSYCH2012 case-cohort for major depressive disorder (MDD), recurrent depressive disorder and schizophrenia (SCZ), we were able to model all individuals directly. As shown in the simulation study, we again outperformed the two genotype-based approaches based on predictive performance of estimated reference-free polygenic risk scores. The gain from using inferred haplo-type clusters of `hapla` was especially large for SCZ, which notably had a significant lower number of cases than the depression disorders. We further compared `hapla` to a reference-based approach using state-of-the-art GWAS summary statistics from European samples of the Psychiatric GWAS Consortium (PGC) for MDD and SCZ. The predictive performance of the reference-based polygenic risk scores for MDD was far below the reference-free polygenic risk scores of all other approaches, while the reference-based polygenic risk scores for SCZ performance almost matched `hapla`. The GWAS summary statistics of the reference-based approach were based on far more cases, especially for SCZ, thus indicating the need for much larger sample sizes to reach the performance of reference-free approaches. However, we further investigated the correlation between the reference-based polygenic risk scores and the estimated principal components in the iPSYCH2012 case-cohort, as it is known to potentially inflate their predictive performance, which was the case for both MDD and SCZ. This means that the polygenic risk scores estimated from the PGC GWASs potentially captures non-genetic effects due to for example uncorrected population structure, thus highlighting further potential bias in relying on external summary statistics. Finally, we demonstrated that `hapla` was more robust in comparison to other approaches, when evaluating the polygenic risk scores for individuals with at least one non-Danish born parent. The well known portability problem affected the reference-based approach to a very large degree, such that it lost all predictive performance for MDD. The results of `hapla` are therefore very promising for real world populations by overcoming some of the inclusion problems that persist in reference-based PRS, as the inferred haplotype clusters were shown to model a structured population better than other standard reference-free approaches, and in general, estimated more predictive polygenic risk scores that can advance the fields of precision medicine and psychiatry.

The standard GREML approach in `GCTA` assumes an infinitesimal prior on the effect sizes and multiple variations have been proposed to either increase the accuracy of heritability estimates [60, 26] or polygenic scores [67, 43]. Our approach can be combined with recent advancements using more complex priors to increase the predictive performance of the reference-free polygenic scores, however, we found it within the scope of our study to solely focus on GREML as a proof of concept with our new data structure, obtaining both unbiased heritability estimates and accurate polygenic scores. Another limitation of our study is that we cluster haplotypes in genomic windows of fixed lengths. In homogeneous populations, it could be more beneficial to use linkage disequilibrium informed blocks as windows [59, 42], as boundaries of the inferred haplotype clusters would reflect frequent breakpoints, tailored to the target population, and potentially eliminate the need for clustering in overlapping windows across multiple window sizes.

With the ability to disentangle genetic information at haplotype level, we hypothesize that our framework can model haplotype-specific effects implicitly through the usage of inferred haplotype clusters instead of SNPs. Thus we can potentially find novel associations, which are not detected in standard SNP-based approaches for structured populations or large-scale multi-population studies. Finally, the data structure of our inferred haplotype clusters opens up exciting future research and extensions to our framework, as the clusters can be used to infer fine-scale population structure and be used in sequential models to infer local ancestry and identity-by-descent tracts as examples. The haplotype cluster medians can as well serve as condensed reference panels for large-scale phasing or imputation problems and potentially mitigate privacy issues.

## Data Availability

Scripts to generate data and reproduce results in the simulation study are available in a public data repository for study replication. Access to the individual-level data of the iPSYCH2012 case-cohort is restricted due to its sensitive nature but the iPSYCH initiative is committed to providing access to the scientific community in accordance with Danish law.

https://github.com/Rosemeis/hapla

## Code availability

The `hapla` software is freely available on GitHub at https://github.com/Rosemeis/hapla under a GNU GPL v3.0 license including code to reproduce the results in the study.

## Acknowledgments

The study was supported by unrestricted grants from the Lundbeck Foundation (R380-2021-1225 and R278-2018-1411) and from the Novo Nordisk Foundation (NNF14CC0001 and NNF23SA0084103). The genotyping of the iPSYCH samples was supported by grants from the Lundbeck Foundation (R102-A9118 and R155-2014-1724), the Stanley Foundation, the Simons Foundation (SFARI 311789), and NIMH (5U01MH094432-02). The funding sources had no role in preparation, review, or approval of the manuscript or the decision to submit for publication.

## Author contributions

J.M. has conceived the study and derived the method. J.M. has implemented the method and performed the analyses. S.R. and M.E.B. have supervised the study. All authors have contributed to the final manuscript.

## Supplementary Material

### Iterative re-clustering of haplotypes

We only include haplotype clusters exceeding a frequency threshold, *δ*, but we perform a post-hoc re-clustering procedure that iteratively eliminates low frequency clusters. The procedure is determined to be converged when the frequency of the haplotype clusters are all above the threshold or when *K* = 2, such that the computational complexity of this procedure is another 𝒪 (*NBK*). The procedure is described in the following algorithm for a single window:

#### Algorithm S1

Iterative re-clustering in `hapla`

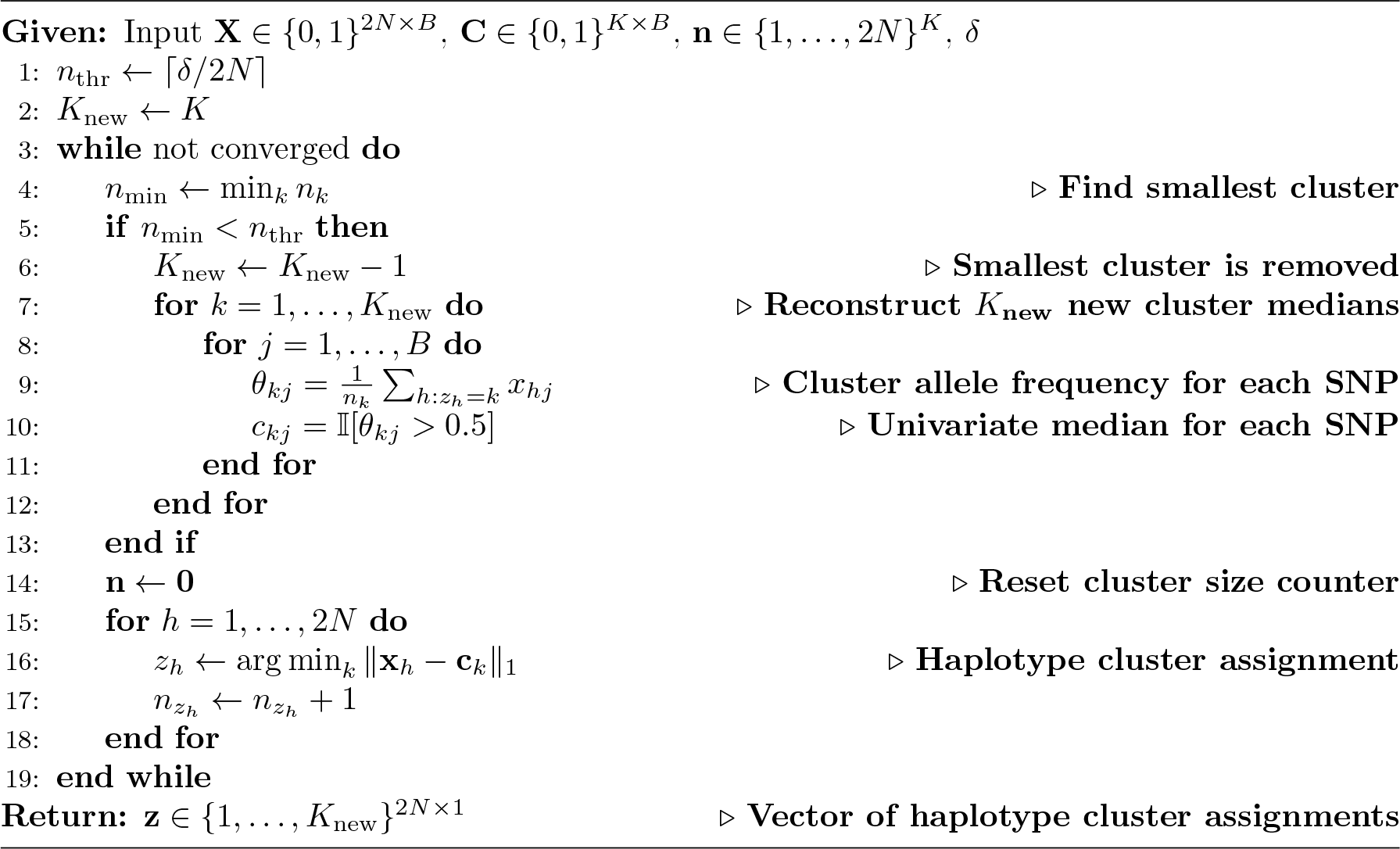

## Supplementary figures

**Figure S1:**
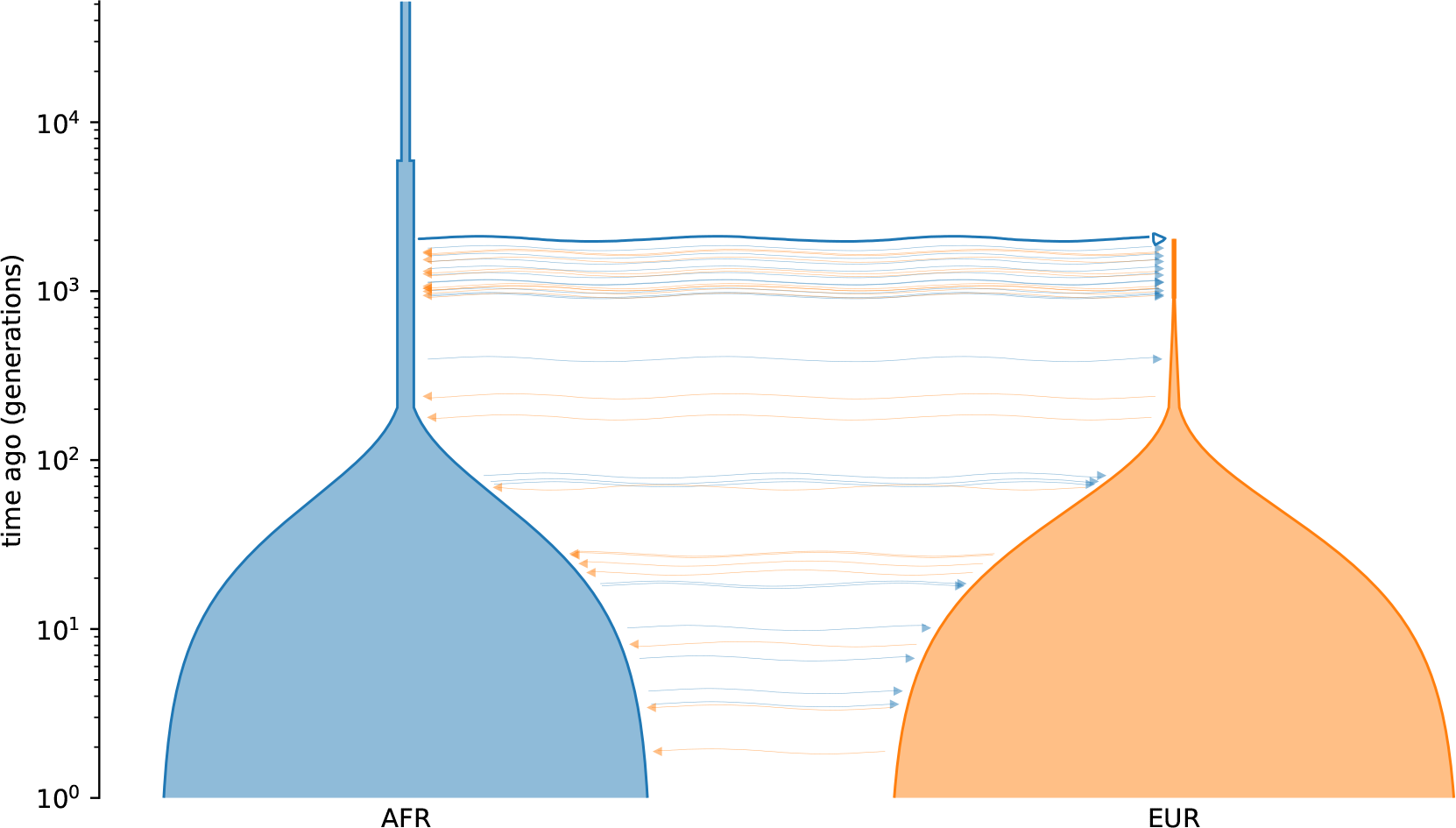
The ‘OutOfAfrica 2T12’ [50, 13] demographic model in `stdpopsim` [1] used for simulations. The figure was generated using the `demesdraw` Python library (https://github.com/grahamgower/demesdraw). 10,000 individuals have been sampled from each of the two populations, AFR and EUR.

**Figure S2:**
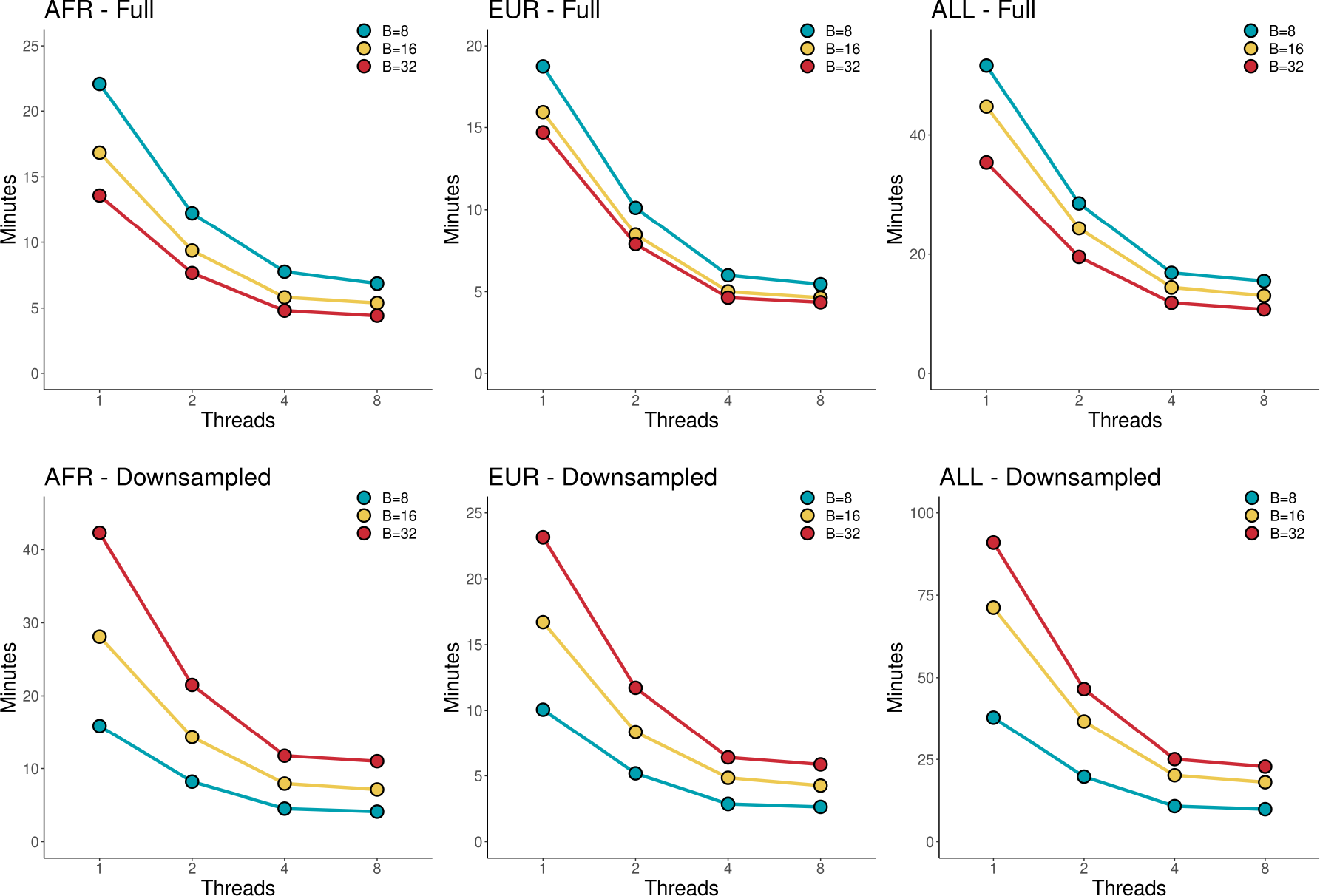
The computational runtimes of haplotype clustering for different window sizes in `hapla`. AFR and EUR consist of 20,000 haplotypes, while ALL consists of 40,000 haplotypes. The top and bottom row shows the computational runtimes for the full and downsampled data, respectively. The tests were performed on a consumer Intel^®^ Core i5-11600 CPU.

**Figure S3:**
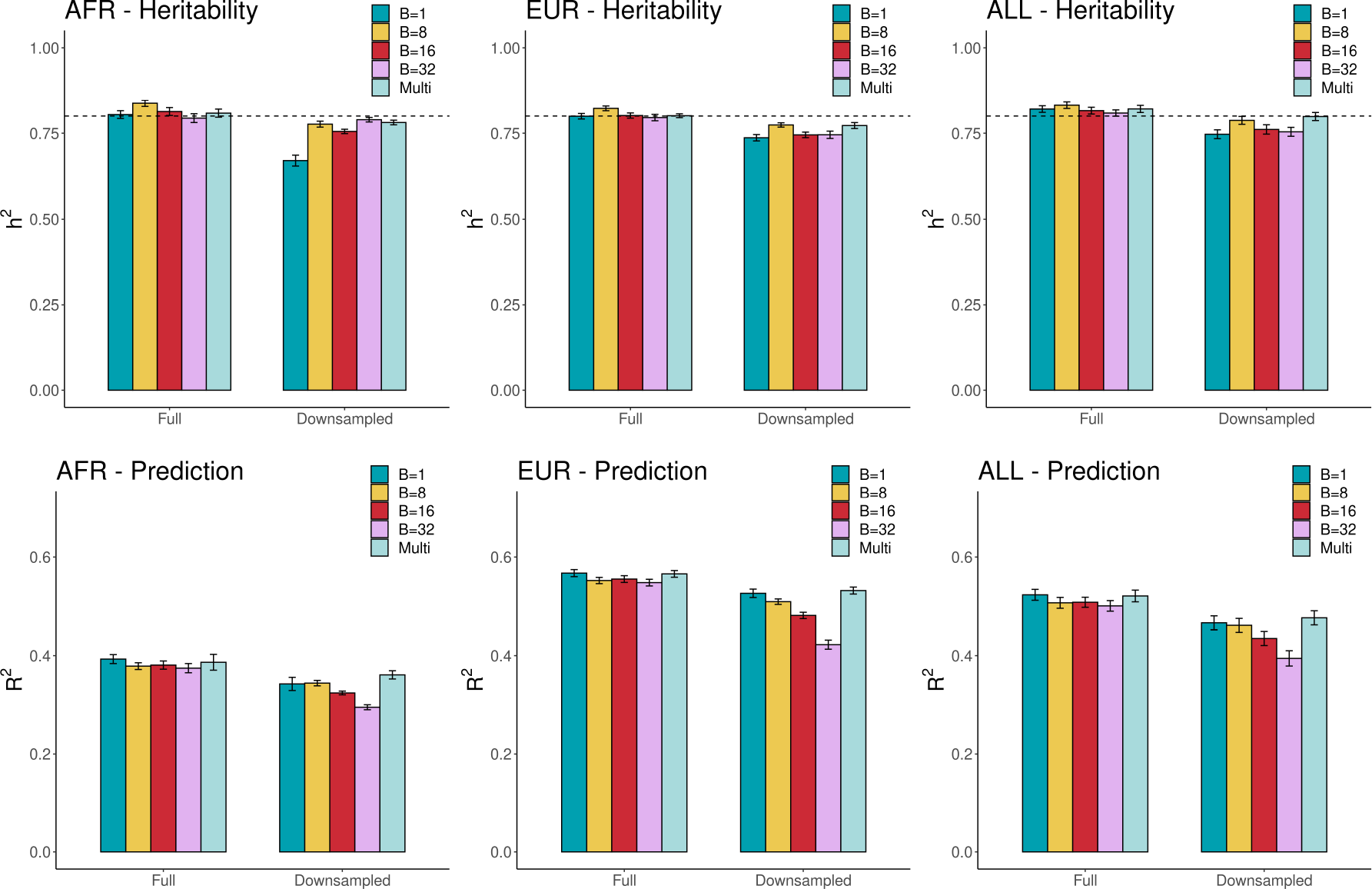
Heritability estimation and polygenic prediction in Scenario 1 across 10 phenotype simulations for different window sizes in `hapla`. “Multi” represents the combined set with all GRMs used in the estimations. Top row shows the SNP heritability estimates using the different GRMs across three data subsets using either full or downsampled data, while the bottom row are the squared correlations between the cvBLUPs and the simulated phenotypes. The true simulated *h*^2^ = 0.8 is displayed with a dashed line and the error bars represent the standard deviation.

**Figure S4:**
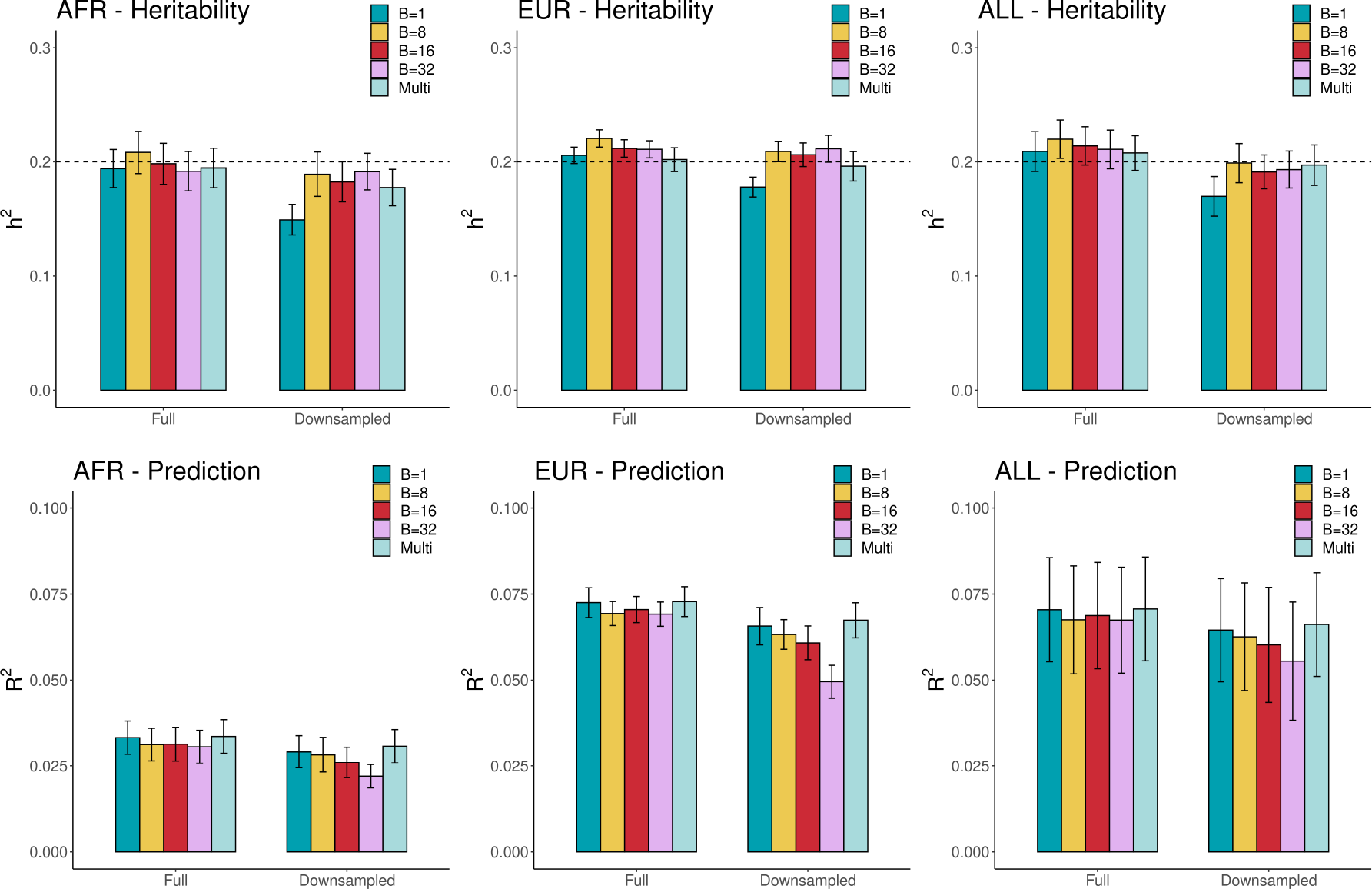
Heritability estimation and polygenic prediction in Scenario 2 across 10 phenotype simulations for different window sizes in `hapla`. “Multi” represents the combined set with all GRMs used in the estimations. Top row shows the SNP heritability estimates using the different GRMs across three data subsets using either full or downsampled data, while the bottom row are the squared correlations between the cvBLUPs and the simulated phenotypes. The true simulated *h*^2^ = 0.2 is displayed with a dashed line and the error bars represent the standard deviation.

**Figure S5:**
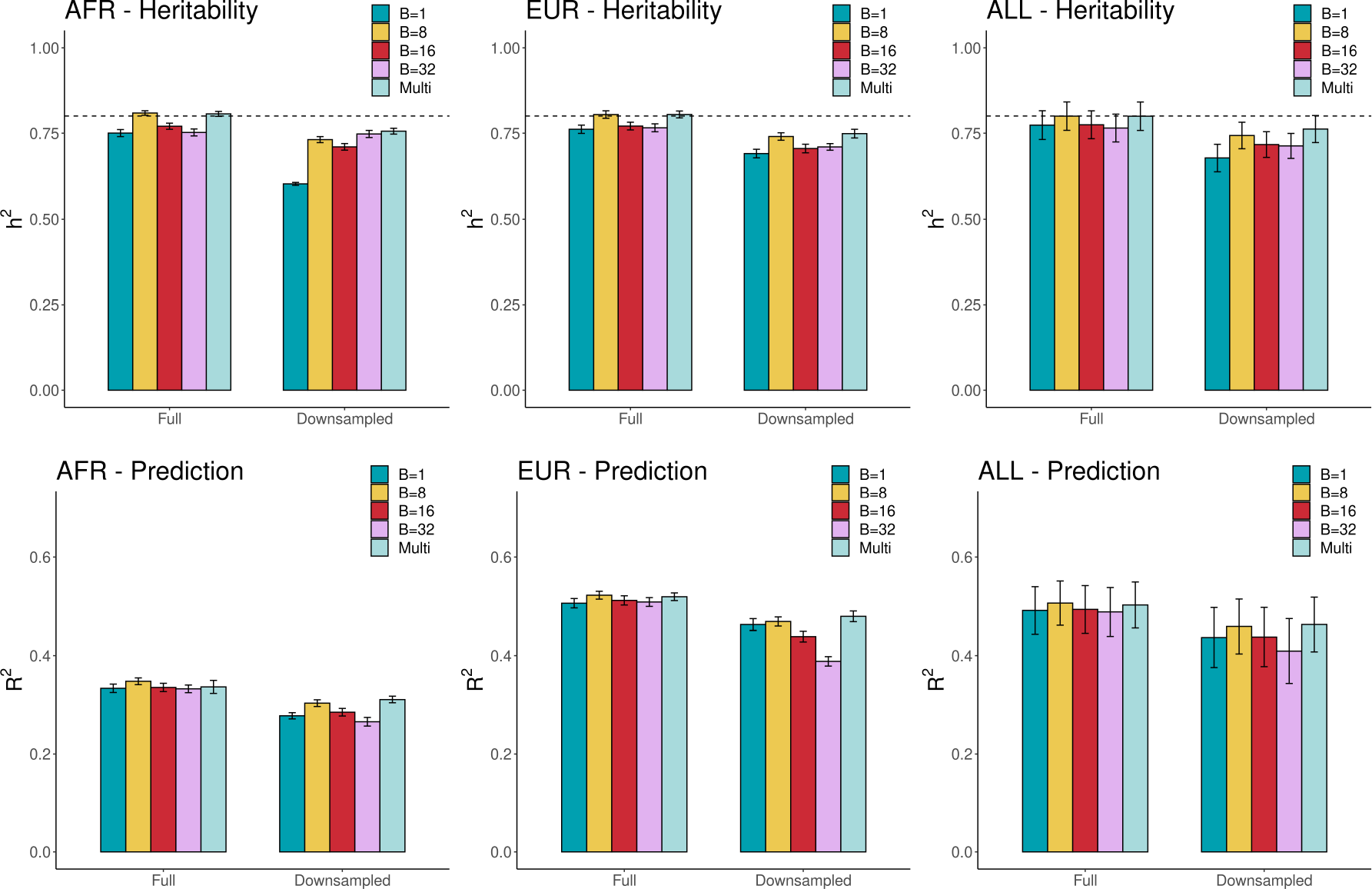
Heritability estimation and polygenic prediction in Scenario 3 (haplotype cluster effects) across 10 phenotype simulations for different window sizes in `hapla`. “Multi” represents the combined set with all GRMs used in the estimations. Top row shows the SNP heritability estimates using the different GRMs across three data subsets using either full or downsampled data, while the bottom row are the squared correlations between the cvBLUPs and the simulated phenotypes. The true simulated *h*^2^ = 0.8 is displayed with a dashed line and the error bars represent the standard deviation.

**Figure S6:**
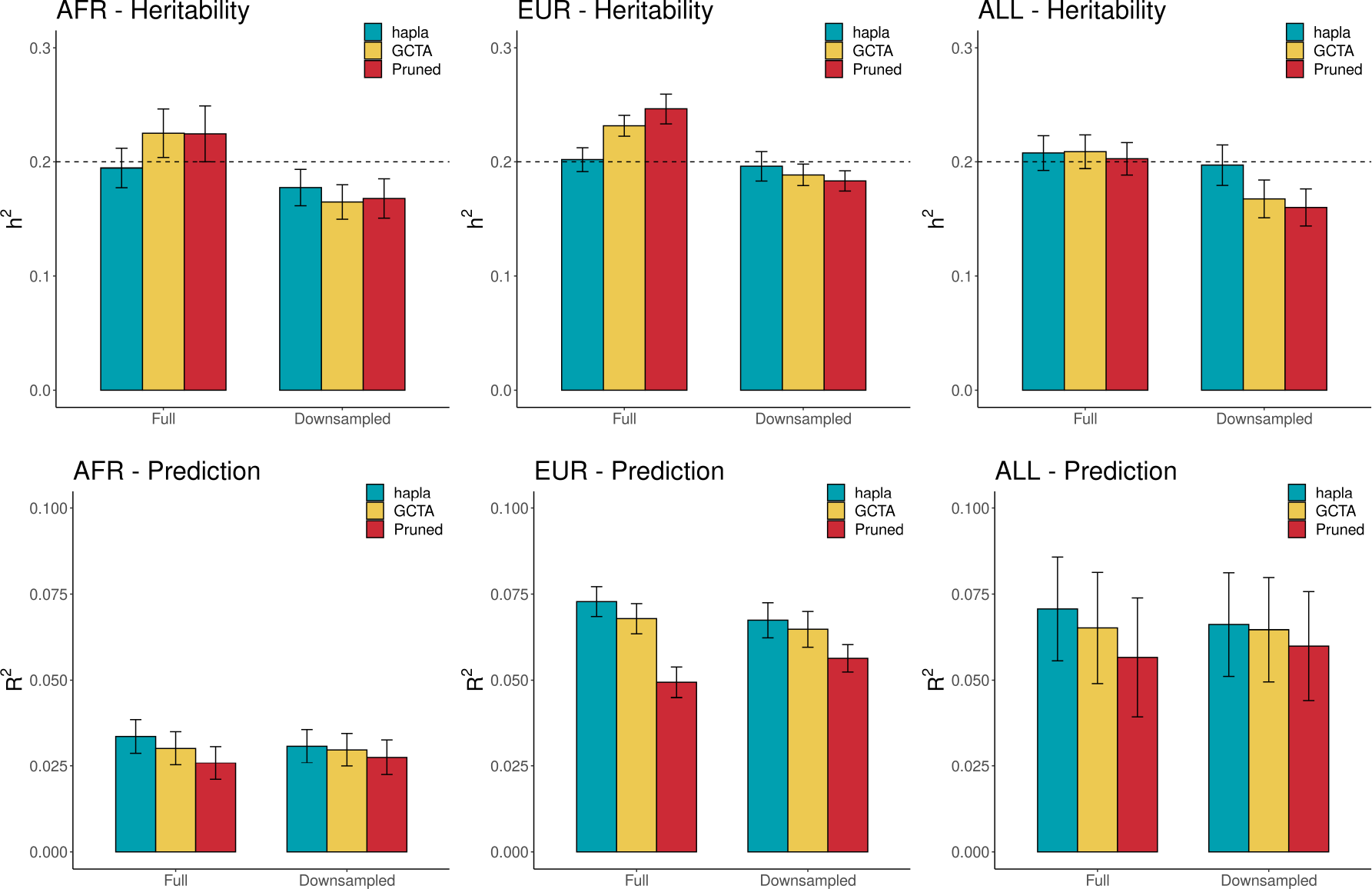
Heritability estimation and polygenic prediction in Scenario 2 across 10 phenotype simulations. Top row shows the SNP heritability estimates using the different GRMs across three data subsets using either full or downsampled data, while the bottom row are the squared correlations between the cvBLUPs and the simulated phenotypes. The true simulated *h*^2^ = 0.2 is displayed with a dashed line and the error bars represent the standard deviation.

**Figure S7:**
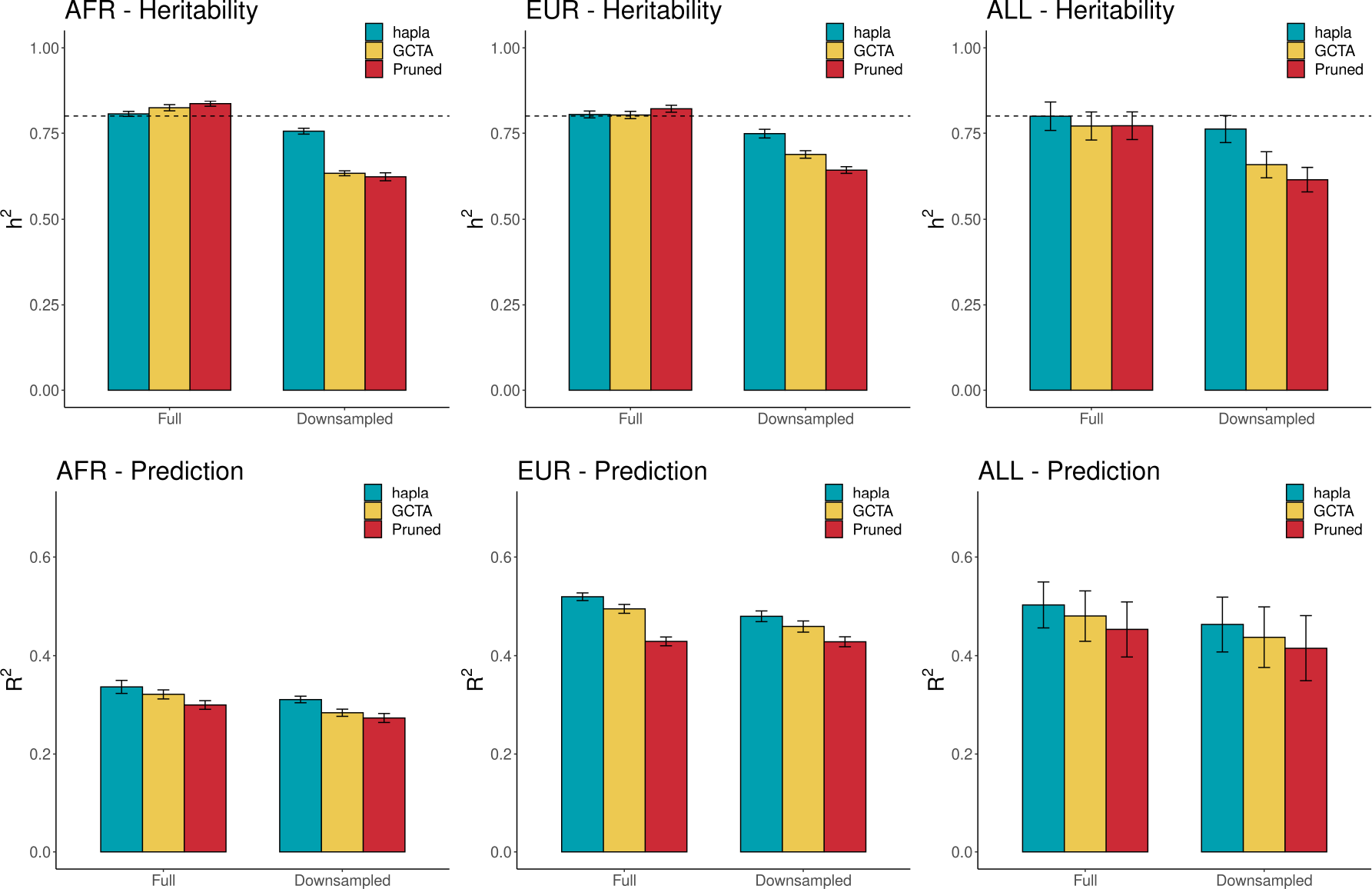
Heritability estimation and polygenic prediction in Scenario 3 (haplotype cluster effects) across 10 phenotype simulations. Top row shows the SNP heritability estimates using the different GRMs across three data subsets using either full or downsampled data, while the bottom row are the squared correlations between the cvBLUPs and the simulated phenotypes. The true simulated *h*^2^ = 0.8 is displayed with a dashed line and the error bars represent the standard deviation.

**Figure S8:**
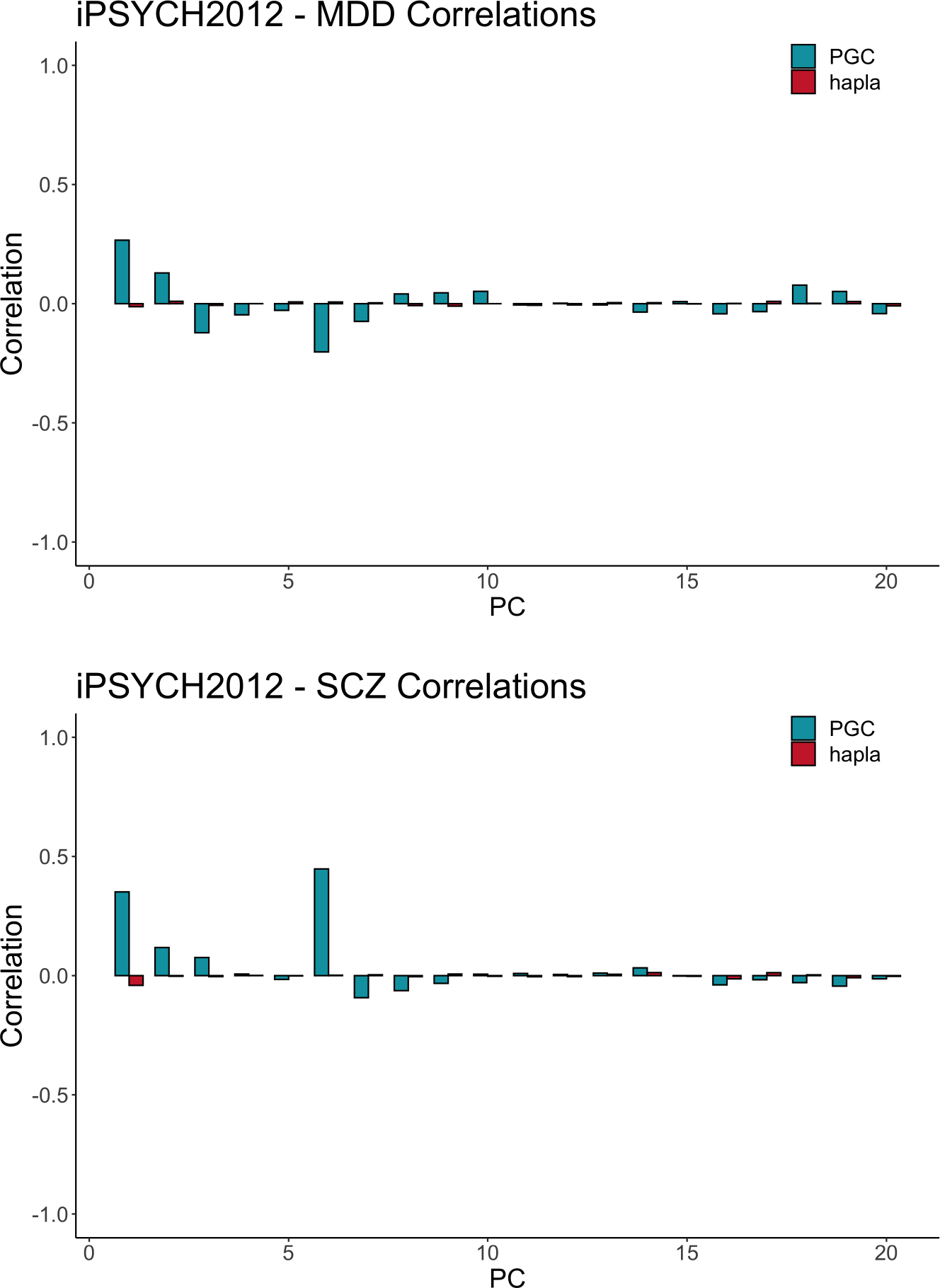
Correlations between the estimated polygenic risk scores and the principal components computed in the iPSYCH2012 case-cohort for major depressive disorder (MDD) and schizophrenia (SCZ) using `PRSice` and `hapla`.

## Supplementary tables

**Table S1:** Overview of the number of SNPs in the simulated datasets. AFR refers to the African ancestry subset, EUR refers to the European ancestry subset and ALL refers to the full set of individuals. The downsampled datasets have been generated by evenly sampling SNPs of the full SNP sets from 10 MAF bins. LD pruning has been performed in `PLINK` (v2.00a) using the flag, “`--indep-pairwise 50 10 0.5`”.

| #SNPs | Full | Downsampled |
| --- | --- | --- |
| **AFR** |  |  |
| No pruning | 425,903 | 42,600 |
| LD pruned | 184,941 | 26,246 |
| **EUR** |  |  |
| No pruning | 223,407 | 22,350 |
| LD Pruned | 58,089 | 10,821 |
| **ALL** |  |  |
| No pruning | 388,746 | 38,880 |
| LD Pruned | 152,089 | 22,059 |

**Table S2:** Overview of the number of windows and inferred haplotype clusters in the simulated datasets using different window sizes for the full and downsampled datasets. AFR refers to the African ancestry subset, EUR refers to the European ancestry subset and ALL refers to the combined set of individuals.

|  | Full |  | Downsampled |  |
| --- | --- | --- | --- | --- |
| **AFR** | #Windows | #Clusters | #Windows | #Clusters |
| $B = 1$ | 425,903 | 851,806 | 42,600 | 85,200 |
| $B = 8$ | 106,473 | 840,728 | 10,649 | 122,589 |
| $B = 16$ | 53,235 | 362,324 | 5,323 | 80,295 |
| $B = 32$ | 26,617 | 194,954 | 2,661 | 75,295 |
| **EUR** | #Windows | #Clusters | #Windows | #Clusters |
| $B = 1$ | 223,407 | 446,814 | 22,350 | 44,700 |
| $B = 8$ | 55,849 | 342,767 | 5,585 | 51,728 |
| $B = 16$ | 27,923 | 153,377 | 2,791 | 32,884 |
| $B = 32$ | 13,961 | 92,298 | 1,395 | 31,173 |
| **ALL** | #Windows | #Clusters | #Windows | #Clusters |
| $B = 1$ | 388,746 | 777,492 | 38,880 | 77,760 |
| $B = 8$ | 97,185 | 749,130 | 9,719 | 109,566 |
| $B = 16$ | 48,591 | 331,527 | 4,859 | 70,806 |
| $B = 32$ | 24,295 | 197,282 | 24,29 | 65,778 |

**Table S3:** Overview of the number of SNPs in the iPSYCH2012 case-cohort datasets. MDD refers to major depressive disorder, REC refers to recurrent depressive disorder and SCZ refers to schizophrenia. LD pruning has been performed in `PLINK` (v2.00a) using the flag, “`--indep-pairwise 50 10 0.5`”.

| **iPSYCH2012** |  |
| --- | --- |
| **MDD** | **#SNPs** |
| No pruning | 5,349,266 |
| LD pruned | 541,038 |
| **REC** | **#SNPs** |
| No pruning | 5,352,358 |
| LD Pruned | 543,287 |
| **SCZ** | **#SNPs** |
| No pruning | 5,356,586 |
| LD Pruned | 544,692 |

**Table S4:** Overview of the number of windows and inferred haplotype clusters in the iPSYCH2012 case-cohort datasets using different window sizes. MDD refers to major depressive disorder, REC refers to recurrent depressive disorder and SCZ refers to schizophrenia.

| **iPSYCH2012** |  |  |
| --- | --- | --- |
| **MDD** | **#Windows** | **#Clusters** |
| $B = 1$ | 5,349,266 | 10,698,532 |
| $B = 8$ | 1,337,280 | 6,573,919 |
| $B = 16$ | 668,614 | 3,320,307 |
| $B = 32$ | 334,290 | 1,994,969 |
| **REC** | **#Windows** | **#Clusters** |
| $B = 1$ | 5,352,358 | 10,704,716 |
| $B = 8$ | 1,338,052 | 8,372,134 |
| $B = 16$ | 669,002 | 4,387,011 |
| $B = 32$ | 334,480 | 2,892,442 |
| **SCZ** | **#Windows** | **#Clusters** |
| $B = 1$ | 5,356,586 | 10,713,172 |
| $B = 8$ | 1,339,104 | 6,595,968 |
| $B = 16$ | 669,526 | 3,332,453 |
| $B = 32$ | 334,744 | 1,992,572 |

**Table S5:** Heritability estimation and polygenic prediction in Scenario 1 across 10 phenotype simulations for different window sizes in `hapla` using either full or downsampled data. “Multi” represents the combined set with all GRMs used in the estimations. The standard deviations are reported in parentheses. The heritability estimated are compared to the true simulated *h*^2^ = 0.8 and the closest estimates are marked in bold, while the highest *R*^2^ values are marked in bold as well for polygenic prediction.

| Scenario 1 | Full |  | Downsampled |  |
| --- | --- | --- | --- | --- |
| AFR | $h^2$ | $R^2$ | $h^2$ | $R^2$ |
| $B = 1$ | **0.8043** (0.0112) | **0.3930** (0.0091) | 0.6702 (0.0159) | 0.3424 (0.0134) |
| $B = 8$ | 0.8373 (0.0093) | 0.3787 (0.0069) | 0.7763 (0.0087) | 0.3440 (0.0055) |
| $B = 16$ | 0.8131 (0.0115) | 0.3808 (0.0084) | 0.7551 (0.0064) | 0.3240 (0.0038) |
| $B = 32$ | 0.7939 (0.0129) | 0.3745 (0.0094) | **0.7893** (0.0066) | 0.2954 (0.0051) |
| Multi | 0.8084 (0.0118) | 0.3865 (0.0161) | 0.7811 (0.0068) | **0.3608** (0.0084) |
| EUR | $h^2$ | $R^2$ | $h^2$ | $R^2$ |
| $B = 1$ | **0.7995** (0.0080) | **0.5677** (0.0072) | 0.7367 (0.0093) | 0.5266 (0.0086) |
| $B = 8$ | 0.8223 (0.0069) | 0.5527 (0.0064) | **0.7740** (0.0063) | 0.5096 (0.0056) |
| $B = 16$ | 0.8014 (0.0076) | 0.5556 (0.0069) | 0.7451 (0.0081) | 0.4815 (0.0064) |
| $B = 32$ | 0.7956 (0.0094) | 0.5484 (0.0068) | 0.7453 (0.0106) | 0.4224 (0.0092) |
| Multi | 0.8009 (0.0055) | 0.5662 (0.0068) | 0.7722 (0.0086) | **0.5322** (0.0071) |
| ALL | $h^2$ | $R^2$ | $h^2$ | $R^2$ |
| $B = 1$ | 0.8204 (0.0096) | **0.5235** (0.0111) | 0.7469 (0.0129) | 0.4665 (0.0142) |
| $B = 8$ | 0.8316 (0.0095) | 0.5072 (0.0111) | 0.7873 (0.0115) | 0.4614 (0.0141) |
| $B = 16$ | 0.8157 (0.0098) | 0.5084 (0.0102) | 0.7609 (0.0136) | 0.4349 (0.0144) |
| $B = 32$ | **0.8087** (0.0092) | 0.5010 (0.0107) | 0.7540 (0.0130) | 0.3944 (0.0155) |
| Multi | 0.8207 (0.0104) | 0.5212 (0.0119) | **0.7986** (0.0117) | **0.4767** (0.0144) |

**Table S6:**
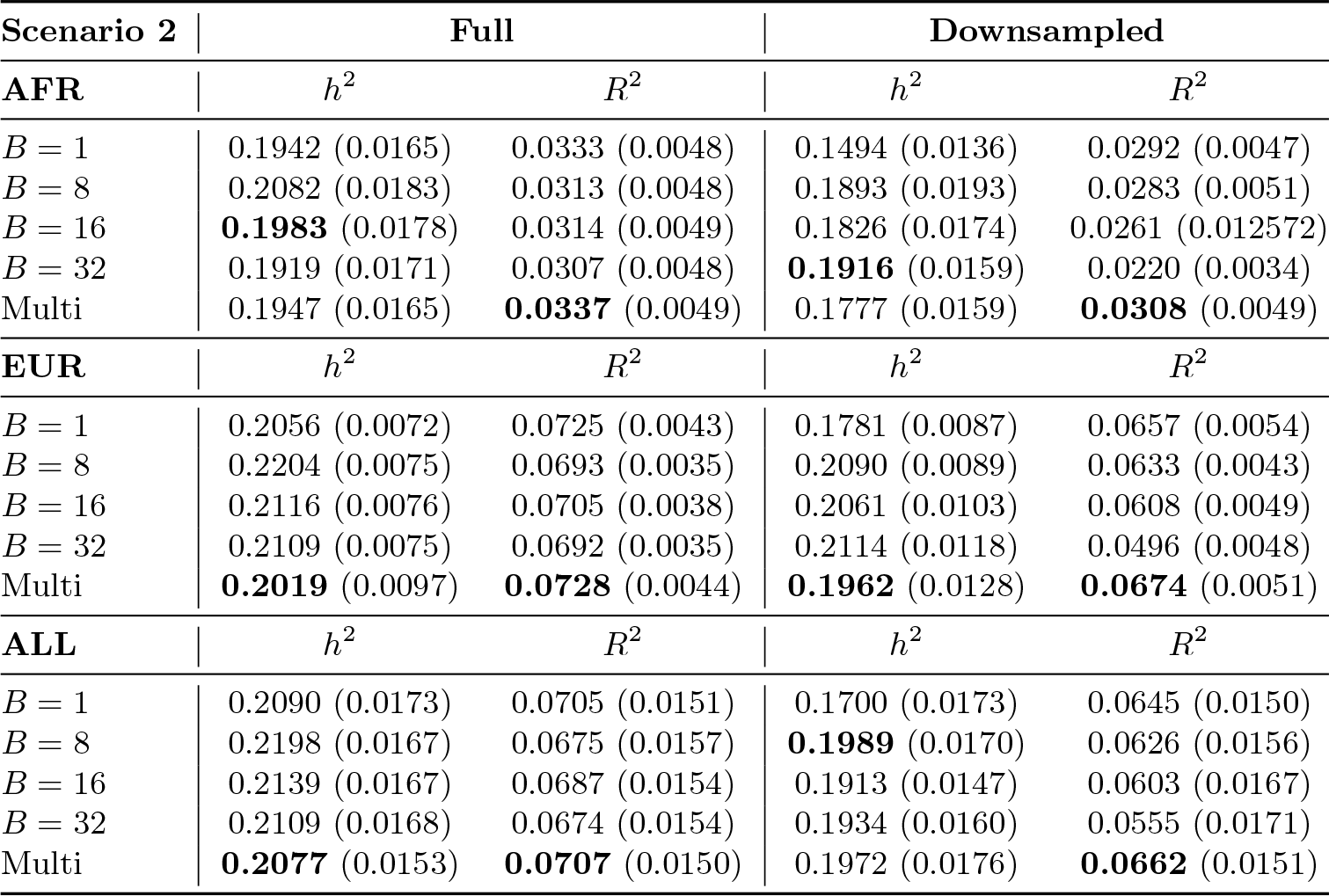
Heritability estimation and polygenic prediction in Scenario 2 across 10 phenotype simulations for different window sizes in `hapla` using either full or downsampled data. “Multi” represents the combined set with all GRMs used in the estimations. The standard deviations are reported in parentheses. The heritability estimated are compared to the true simulated *h*^2^ = 0.2 and the closest estimates are marked in bold, while the highest *R*^2^ values are marked in bold as well for polygenic prediction.

| Scenario 2 | Full |  | Downsampled |  |
| --- | --- | --- | --- | --- |
| AFR | $h^2$ | $R^2$ | $h^2$ | $R^2$ |
| $B = 1$ | 0.1942 (0.0165) | 0.0333 (0.0048) | 0.1494 (0.0136) | 0.0292 (0.0047) |
| $B = 8$ | 0.2082 (0.0183) | 0.0313 (0.0048) | 0.1893 (0.0193) | 0.0283 (0.0051) |
| $B = 16$ | **0.1983** (0.0178) | 0.0314 (0.0049) | 0.1826 (0.0174) | 0.0261 (0.012572) |
| $B = 32$ | 0.1919 (0.0171) | 0.0307 (0.0048) | **0.1916** (0.0159) | 0.0220 (0.0034) |
| Multi | 0.1947 (0.0165) | **0.0337** (0.0049) | 0.1777 (0.0159) | **0.0308** (0.0049) |
| EUR | $h^2$ | $R^2$ | $h^2$ | $R^2$ |
| $B = 1$ | 0.2056 (0.0072) | 0.0725 (0.0043) | 0.1781 (0.0087) | 0.0657 (0.0054) |
| $B = 8$ | 0.2204 (0.0075) | 0.0693 (0.0035) | 0.2090 (0.0089) | 0.0633 (0.0043) |
| $B = 16$ | 0.2116 (0.0076) | 0.0705 (0.0038) | 0.2061 (0.0103) | 0.0608 (0.0049) |
| $B = 32$ | 0.2109 (0.0075) | 0.0692 (0.0035) | 0.2114 (0.0118) | 0.0496 (0.0048) |
| Multi | **0.2019** (0.0097) | **0.0728** (0.0044) | **0.1962** (0.0128) | **0.0674** (0.0051) |
| ALL | $h^2$ | $R^2$ | $h^2$ | $R^2$ |
| $B = 1$ | 0.2090 (0.0173) | 0.0705 (0.0151) | 0.1700 (0.0173) | 0.0645 (0.0150) |
| $B = 8$ | 0.2198 (0.0167) | 0.0675 (0.0157) | **0.1989** (0.0170) | 0.0626 (0.0156) |
| $B = 16$ | 0.2139 (0.0167) | 0.0687 (0.0154) | 0.1913 (0.0147) | 0.0603 (0.0167) |
| $B = 32$ | 0.2109 (0.0168) | 0.0674 (0.0154) | 0.1934 (0.0160) | 0.0555 (0.0171) |
| Multi | **0.2077** (0.0153) | **0.0707** (0.0150) | 0.1972 (0.0176) | **0.0662** (0.0151) |

**Table S7:** Heritability estimation and polygenic prediction in Scenario 3 (haplotype cluster effects) across 10 phenotype simulations for different window sizes in `hapla` using either full or downsampled data. “Multi” represents the combined set with all GRMs used in the estimations. The standard deviations are reported in parentheses. The heritability estimated are compared to the true simulated *h*^2^ = 0.8 and the closest estimates are marked in bold, while the highest *R*^2^ values are marked in bold as well for polygenic prediction.

| Scenario 3 | Full |  | Downsampled |  |
| --- | --- | --- | --- | --- |
| AFR | $h^2$ | $R^2$ | $h^2$ | $R^2$ |
| $B = 1$ | 0.7503 (0.0102) | 0.3335 (0.0086) | 0.6032 (0.0042) | 0.2780 (0.0063) |
| $B = 8$ | 0.8087 (0.0065) | **0.3478** (0.0069) | 0.7313 (0.0088) | 0.3037 (0.0068) |
| $B = 16$ | 0.7702 (0.0089) | 0.3352 (0.0085) | 0.7101 (0.0096) | 0.2853 (0.0077) |
| $B = 32$ | 0.7521 (0.0101) | 0.3324 (0.0078) | 0.7477 (0.0103) | 0.2658 (0.0088) |
| Multi | **0.8062** (0.0073) | 0.3363 (0.0133) | **0.7558** (0.0085) | **0.3110** (0.0064) |
| EUR | $h^2$ | $R^2$ | $h^2$ | $R^2$ |
| $B = 1$ | 0.7614 (0.0121) | 0.5064 (0.0095) | 0.6906 (0.0125) | 0.4631 (0.0119) |
| $B = 8$ | **0.8043** (0.0109) | **0.5227** (0.0080) | 0.7403 (0.0109) | 0.4692 (0.0093) |
| $B = 16$ | 0.7707 (0.0112) | 0.5122 (0.0094) | 0.7054 (0.0125) | 0.4388 (0.0110) |
| $B = 32$ | 0.7656 (0.0115) | 0.5089 (0.0089) | 0.7100 (0.0096) | 0.3883 (0.0096) |
| Multi | 0.8045 (0.0100) | 0.5196 (0.0078) | **0.7487** (0.0127) | **0.4797** (0.0109) |
| ALL | $h^2$ | $R^2$ | $h^2$ | $R^2$ |
| $B = 1$ | 0.7735 (0.0418) | 0.4918 (0.0482) | 0.6780 (0.0397) | 0.4368 (0.0611) |
| $B = 8$ | **0.7996** (0.0414) | **0.5067** (0.0450) | 0.7433 (0.0386) | 0.4592 (0.0558) |
| $B = 16$ | 0.7745 (0.0406) | 0.4939 (0.0484) | 0.7169 (0.0374) | 0.4378 (0.0603) |
| $B = 32$ | 0.7650 (0.0405) | 0.4887 (0.0497) | 0.7129 (0.0363) | 0.4092 (0.0660) |
| Multi | 0.7994 (0.0414) | 0.5029 (0.0468) | **0.7620** (0.0394) | **0.4632** (0.0556) |

**Table S8:** Heritability estimation and polygenic prediction in Scenario 1 across 10 phenotype simulations using either full or downsampled data. The standard deviations are reported in parentheses. The heritability estimated are compared to the true simulated *h*^2^ = 0.8 and the closest estimates are marked in bold, while the highest *R*^2^ values are marked in bold as well for polygenic prediction.

| Scenario 1 | Full |  | Downsampled |  |
| --- | --- | --- | --- | --- |
| AFR | $h^2$ | $R^2$ | $h^2$ | $R^2$ |
| hapla | **0.8084** (0.0118) | **0.3865** (0.0161) | **0.7811** (0.0068) | **0.3608** (0.0084) |
| GCTA | 0.8672 (0.0098) | 0.3638 (0.0071) | 0.6961 (0.0144) | 0.3447 (0.0120) |
| Pruned | 0.8634 (0.0094) | 0.3231 (0.0073) | 0.6791 (0.0149) | 0.3218 (0.0124) |
| EUR | $h^2$ | $R^2$ | $h^2$ | $R^2$ |
| hapla | **0.8009** (0.0055) | **0.5662** (0.0068) | **0.7722** (0.0086) | **0.5322** (0.0071) |
| GCTA | 0.8328 (0.0072) | 0.5453 (0.0085) | 0.7305 (0.0082) | 0.5181 (0.0085) |
| Pruned | 0.8338 (0.0070) | 0.4465 (0.0094) | 0.6831 (0.0093) | 0.4754 (0.0096) |
| ALL | $h^2$ | $R^2$ | $h^2$ | $R^2$ |
| hapla | 0.8207 (0.0104) | **0.5212** (0.0119) | **0.7986** (0.0117) | **0.4767** (0.0144) |
| GCTA | 0.8117 (0.0091) | 0.4945 (0.0113) | 0.7234 (0.0118) | 0.4614 (0.0134) |
| Pruned | **0.8043** (0.0104) | 0.4398 (0.0114) | 0.6776 (0.0087) | 0.4243 (0.0115) |

**Table S9:** Heritability estimation and polygenic prediction in Scenario 2 across 10 phenotype simulations using either full or downsampled data. The standard deviations are reported in parentheses. The heritability estimated are compared to the true simulated *h*^2^ = 0.2 and the closest estimates are marked in bold, while the highest *R*^2^ values are marked in bold as well for polygenic prediction.

| Scenario 2 | Full |  | Downsampled |  |
| --- | --- | --- | --- | --- |
| AFR | $h^2$ | $R^2$ | $h^2$ | $R^2$ |
| hapla | **0.1947** (0.0165) | **0.0337** (0.0049) | **0.1777** (0.0159) | **0.0308** (0.0049) |
| GCTA | 0.2250 (0.0212) | 0.0302 (0.0049) | 0.1651 (0.0151) | 0.0298 (0.0048) |
| Pruned | 0.2244 (0.0244) | 0.0259 (0.0048) | 0.1682 (0.0172) | 0.0276 (0.0051) |
| EUR | $h^2$ | $R^2$ | $h^2$ | $R^2$ |
| hapla | **0.2019** (0.0097) | **0.0728** (0.0044) | **0.1962** (0.0128) | **0.0674** (0.0051) |
| GCTA | 0.2315 (0.0091) | 0.0679 (0.0044) | 0.1887 (0.0093) | 0.0648 (0.0052) |
| Pruned | 0.2463 (0.0132) | 0.0494 (0.0044) | 0.1835 (0.0089) | 0.0564 (0.0040) |
| ALL | $h^2$ | $R^2$ | $h^2$ | $R^2$ |
| hapla | 0.2077 (0.0153) | **0.0707** (0.0150) | **0.1972** (0.0176) | **0.0662** (0.0151) |
| GCTA | 0.2089 (0.0147) | 0.0652 (0.0162) | 0.1678 (0.0165) | 0.0647 (0.0152) |
| Pruned | **0.2027** (0.0141) | 0.0566 (0.0172) | 0.1603 (0.0162) | 0.0599 (0.0158) |

**Table S10:**
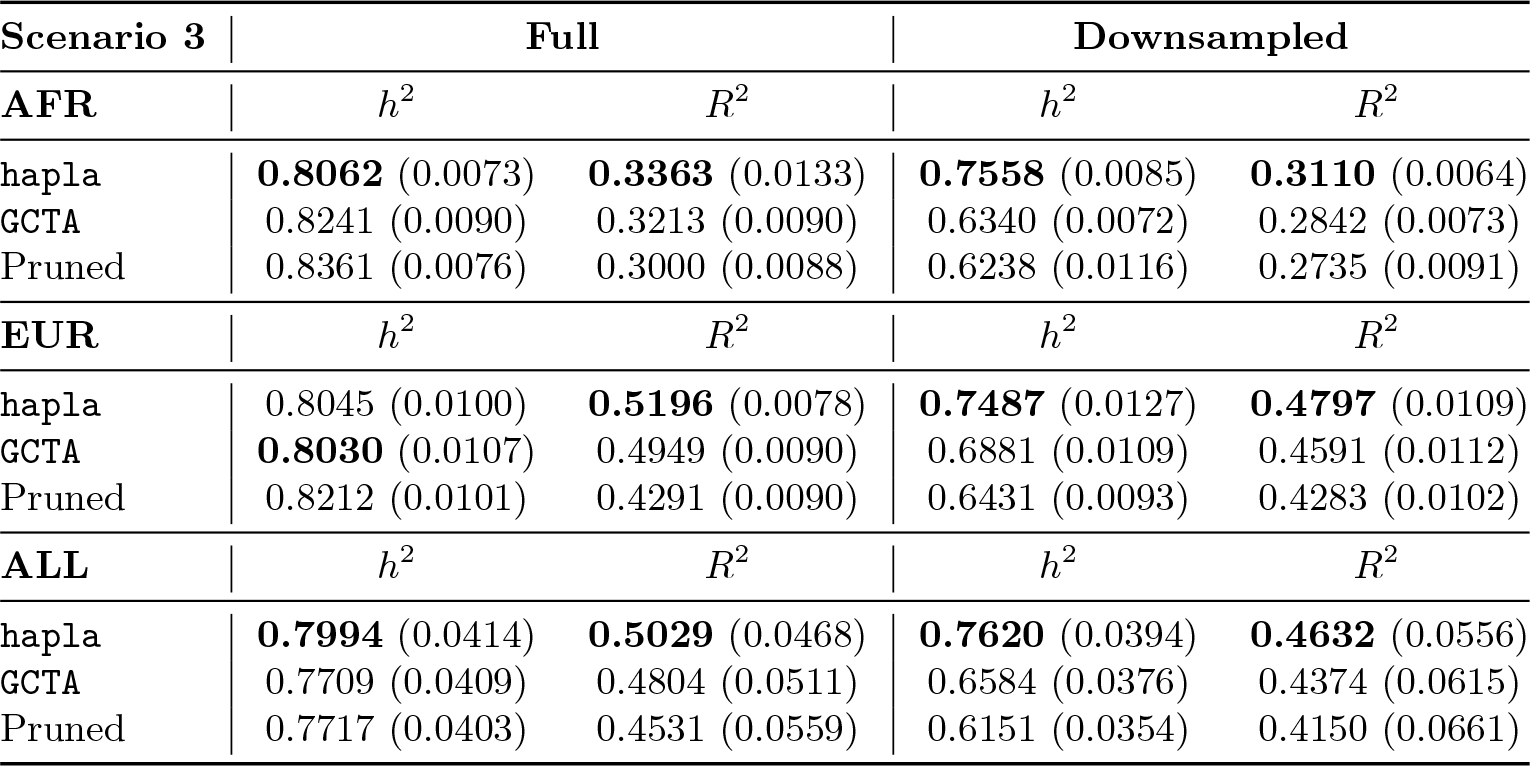
Heritability estimation and polygenic prediction in Scenario 3 (haplotype cluster effects) across 10 phenotype simulations using either full or downsampled data. The standard deviations are reported in parentheses. The heritability estimated are compared to the true simulated *h*^2^ = 0.8 and the closest estimates are marked in bold, while the highest *R*^2^ values are marked in bold as well for polygenic prediction.

**Table S11:** Description of PGC GWAS summary statistics [15, 53] excluding iPSYCH case-cohorts and the performances of the reference-based polygenic risk scores estimated in `PRSice`. #SNPs indicates the number of overlapping SNPs between the iPSYCH2012 case-cohort and the summary statistics, while #SNPs C+T indicates the final number of SNPs used for the polygenic risk scores estimation after clumping and thresholding, given the listed threshold.

| iPSYCH2012 - PGC GWAS summary statistics |  |  |  |  |  |  |
| --- | --- | --- | --- | --- | --- | --- |
| | $N_{\text{cases}}$ | $N_{\text{controls}}$ | #SNPs | #SNPs C+T | Threshold | $R_N^2$ |
| MDD (2018) | 49,982 | 136,871 | 4,457,161 | 18,785 | 0.0446 | 0.0047 |
| SCZ (2022) | 64,952 | 88,856 | 4,430,890 | 19,401 | 0.0263 | 0.0121 |

**Table S12:** Predictive performance of polygenic risk scores in subsets of the iPSYCH2012 case-cohort. “Full sample” refers to the full sample set of individuals in the iPSYCH2012 case-cohort, “Danish born parents” refers to the subset of individuals with both parents being born in Denmark, while “Non-Danish born parent” refers to the subset of individuals with at least one parent born outside Denmark. “PGC” represents the reference-based polygenic risk scores from PGC GWAS summary statistics using PRSice. The predictive performances are reported using Nagelkerke’s pseudo 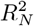, and the highest values are marked in bold.

| iPSYCH2012 - Predictive performances in subsets |  |  |  |  |  |  |
| --- | --- | --- | --- | --- | --- | --- |
| MDD | $N_{\text{cases}}$ | $N_{\text{controls}}$ | hapla | GCTA | Pruned | PGC |
| Full sample | 20,620 | 25,228 | **0.0241** | 0.0225 | 0.0197 | 0.0047 |
| Danish born parents | 18,326 | 21,530 | **0.0246** | 0.0236 | 0.0206 | 0.0094 |
| Non-Danish born parent | 2,294 | 3,698 | **0.0226** | 0.0191 | 0.0170 | 0.0002 |
| REC | $N_{\text{cases}}$ | $N_{\text{controls}}$ | hapla | GCTA | Pruned | PGC |
| Full sample | 7,128 | 25,840 | **0.0107** | 0.0095 | 0.0081 | - |
| Danish born parents | 6,426 | 22,078 | **0.0108** | 0.0098 | 0.0082 | - |
| Non-Danish born parent | 702 | 3,762 | **0.0103** | 0.0089 | 0.0086 | - |
| SCZ | $N_{\text{cases}}$ | $N_{\text{controls}}$ | hapla | GCTA | Pruned | PGC |
| Full sample | 3,211 | 25,994 | **0.0122** | 0.0109 | 0.0094 | 0.0121 |
| Danish born parents | 2,651 | 22,230 | **0.0122** | 0.0110 | 0.0095 | 0.0115 |
| Non-Danish born parent | 560 | 3,764 | **0.0124** | 0.0090 | 0.0076 | 0.0079 |

